# Addiction Potential of Combustible Menthol Cigarette Alternatives: A Randomized Crossover Trial

**DOI:** 10.1101/2022.06.16.22276504

**Authors:** Theodore L. Wagener, Toral Mehta, Alice Hinton, Jonathan Schulz, Tyler Erath, Jennifer W. Tidey, Andrea C. Villanti

## Abstract

**Introduction:** The FDA announced its intention to issue proposed product standards banning menthol as a characterizing flavor in cigarettes and cigars. The public health benefits of these product standards may be attenuated by the role of plausible substitutes in the marketplace. Therefore, the present study examined the addiction potential of plausible combustible menthol alternatives compared to usual brand menthol cigarettes (UBMC).

**Methods:** Eighty adult menthol cigarette smokers completed four visits, smoking their UBMC at the first session and 3 menthol cigarette alternatives in random order at the subsequent visits: 1) a pre-assembled menthol roll-your-own cigarette using menthol pipe tobacco and mentholated cigarette tube (mRYO), 2) a menthol filtered little cigar (mFLC), and 3) a non-menthol cigarette (NMC). Measures of smoking topography, exhaled carbon monoxide (CO), craving and withdrawal, subjective effects, and behavioral economic demand indices were assessed.

**Results:** Compared to UBMC, menthol cigarette alternatives resulted in similar topography and CO exposure but significantly lower levels of positive subjective experience and behavioral economic demand indices. Among the alternative products, participants reported the highest level of positive subjective experience and higher demand for mRYO, compared with mFLC and NMC. Similarly, participants were significantly more likely to want to try again, purchase, and use the mRYO product regularly compared with mFLC and NMC.

**Conclusions and Relevance:** mRYO cigarettes were the most highly rated cigarette alternative among study products, suggesting their potential appeal as a menthol cigarette substitute and needed inclusion of menthol pipe tobacco and cigarette tubes in FDA’s proposed ban.

**What is already known on this topic:** Menthol cigarettes are associated with increased smoking initiation, higher nicotine dependence, and decreased adult cessation, particularly among vulnerable populations. To address this public health issue, the FDA announced in April 2021 its intention to issue product standards banning menthol as a characterizing flavor in both cigarettes and cigars within a year. However, the public health benefits of these product standards may be attenuated by the role of plausible substitutes available in the marketplace

**What this study adds:** In this randomized cross-over design study that included 80 adult menthol cigarette smokers, each of the alternative products demonstrated the ability to significantly reduce nicotine craving and withdrawal symptoms, but the combination of mentholated pipe tobacco and tubes in a menthol roll-your-own cigarette, resulted in the highest behavioral economic demand and positive subjective experience.

**How this study may affect policy:** To maximize the benefits of a menthol cigarette ban, restrictions should extend to plausible substitutes, particularly menthol pipe tobacco and cigarette tubes.

## INTRODUCTION

The decreasing prevalence of cigarette smoking in the U.S.^1^ has been driven by decreases in non-menthol cigarette use.^2 3^ In contrast, menthol cigarette consumption has remained largely stable since 2000, resulting in a more than 10% increase in menthol cigarette market share over the past two decades^2^ and reaching 37% market share in 2020.^4^ Menthol cigarettes are associated with increased smoking initiation and progression to regular use,^5 6^ higher nicotine dependence, and decreased adult cessation,^7^ particularly among Black people who smoke cigarettes.^8 9^ People who smoke menthol cigarettes are also more likely to be of low socioeconomic status, female, Black or Hispanic, and identify as LGBT compared to non-menthol smokers.^10 11^ Menthol cigarette smoking is estimated to have caused 10.1 million extra smokers, 3 million life years lost, and 378,000 premature deaths between 1980 and 2018,^12^ with disproportionate harms experienced by Black Americans.^13^

The 2009 Family Smoking Prevention and Tobacco Control Act granted the US FDA broad authority to regulate tobacco products, leading to bans of flavored cigarettes, excluding menthol, and some flavored e-cigarette devices. In April 2021, the FDA announced its intention to issue product standards banning menthol as a characterizing flavor in both cigarettes and cigars within a year.^14^ While evidence from systematic reviews,^7 15-17^ evaluations of Ontario’s menthol cigarette ban,^18-20^ and simulation studies strongly support the likely positive public health impact of a menthol ban on cigarette and cigars,^21-23^ experimental evidence is also needed to bolster these findings to withstand tobacco industry lawsuits.

Even with regulation, tobacco companies frequently exploit regulatory loopholes to maintain sales of their products.^24 25^ Djarum, for example, launched clove filtered cigars in the U.S. in anticipation of the 2009 ban on flavored cigarettes and sales of clove filtered cigars increased by more than 1400% between 2009 and 2012.^26^ Similarly, following the 2009 Children’s Health Insurance Program Reauthorization Act (CHIPRA), which levied large increases in federal excise tax rates on cigarettes, cigars, and roll-your-own (RYO) tobacco, tobacco companies repackaged and labeled RYO tobacco as pipe tobacco to avoid these policies.^27^ As a result, while the market share of pipe tobacco declined from 30.4% in 2002 to 13.6% in 2008, it increased significantly to 89.6% in 2012.^27^ This reflects an increase of approximately 25.49 million pounds of loose tobacco (i.e., RYO and pipe tobacco) sold per year from 2002 to 2012.^27^ These examples highlight how the potential public health benefits of regulation, such as a product standard, can be attenuated and suggest that estimating the impact of a potential ban on menthol in cigarettes requires accounting for likely substitutes in the marketplace that may also need to be restricted to effectively protect public health. Current tobacco products, including menthol filtered little cigars, menthol pipe tobacco and cigarette tubes, and non-menthol cigarettes, are relevant targets.^28^

The purpose of the present study was to assess the addiction potential of other plausible combustible menthol cigarette alternatives in adults who smoke menthol cigarettes by examining the impact of these alternatives on subjective effects, behavioral economic demand indices, smoking topography, and resultant toxicant exposure compared to the participants’ usual brand menthol cigarette (UBMC). We hypothesized that compared to UBMC, alternatives would result in similar smoking topography and carbon monoxide (CO) exposure, but fewer positive subjective effects and lower demand.

## METHODS

### Setting and Participants

Menthol cigarette smokers from the Columbus, Ohio metropolitan area were recruited via internet advertisements, flyers, and word-of-mouth advertising from January 2020 to August 2021. Potential participants were screened for eligibility via an online questionnaire and then over the telephone. Eligibility criteria included (1) current menthol cigarette smoker (>90% menthol cigarette use; ≥5 cigarettes per day) for at least the past 6 months; (2) between 21 and 50 years old; (3) willing to abstain from tobacco, nicotine and marijuana use for at least 12 hours prior to each of the study visits; (4) access to a smartphone or email and (5) read and speak English. Exclusionary criteria included (1) self-reported diagnosis of lung disease; (2) cardiac event or distress within the past 3 months; (3) pregnancy, breastfeeding, or planning to become pregnant; (4) use of other tobacco products (e.g., e-cigarette, cigar, etc.) >5 days in the past month; (5) currently using one of the study products; (6) any reported use of illicit drugs (other than marijuana) during the last 30 days; and (7) currently engaging in smoking cessation treatment. All participants provided written informed consent.

### Procedure

Using an in-laboratory and outpatient mixed design, participants completed a three-phase study lasting approximately three weeks. In phase 1, participants completed 4 smoking session visits, smoking their UBMC or one of the 3 menthol cigarette alternatives at each visit. Each visit was separated by a 48-hour washout period. This phase used multiple methods of assessing addiction potential in a lab-based setting, including measurements of drug self-administration; suppression of craving and withdrawal; measures of drug liking; and behavioral economic measures.^29^ In phase 2, participants were instructed to completely substitute their preferred product from phase 1 for their UBMC for one week and complete daily assessments of their use behavior. In phase 3, participants completed a final in-lab visit to assess the substitutability of their preferred product, under simulated ban conditions using a progressive ratio task. We report the results of phase 1 below.

All participants completed sociodemographic measures and provided their tobacco use history including their years of smoking, usual cigarette brand, smoking frequency and quantity, number and recency of previous quit attempts, and their level of cigarette dependence (Fagerstrom Test for Nicotine Dependence^30^). Then, over four visits, participants completed standardized smoking sessions, smoking a different product each session. During each session, a research assistant with a stopwatch instructed participants to take a puff every 30 s, resulting in 10 puffs during the first 5 min. Participants sampled their UBMC during the first session; the order of subsequent products was randomized at the time of enrollment from a prespecified block randomization table with blocks of size 6. Products included a pre-assembled (by study staff), machine-injected roll-your-own cigarette using a mentholated cigarette tube and mentholated pipe tobacco (mRYO; OHM menthol pipe tobacco, hot rod tubes - menthol king size), a mentholated filtered little cigar (mFLC; Cheyenne 100’s menthol) and a non-menthol cigarette (NMC; Newport Non-Menthol Red). All products were provided in plain boxes without brand or identifying information. Brand name was present, however, on the NMC filter wrapper. Participants abstained from smoking (biochemically confirmed exhaled carbon monoxide ≤ 10ppm), nicotine, and marijuana for 12 hours before the sessions.

During the smoking session, participant puffing topography was collected including average flow rate, inter-puff interval, puff volume, puff duration, maximum puff volume, total puff time, total smoking time, and total inhaled volume. Measures of cigarette craving (Tiffany-Drobes Questionnaire of Smoking Urges [QSU]: Brief Form^31^) and withdrawal (Minnesota Nicotine Withdrawal Scale [MNWS]^32^) were also administered immediately before (0) and after the smoking session (5 min), and at 15, 30, 60, and 90 min. The QSU is a 10-item self-report measure with items rated from 1 (strongly disagree) to 7 (strongly agree). Items have been shown to load on two factors— ‘desire to smoke’ and ‘anticipated relief from withdrawal’. For the MNWS, completed the 15-item version but for analysis we used the 9-item version assessing the Diagnostic and Statistical Manual of Mental Disorders (DSM-5) symptoms for Tobacco Withdrawal and the “craving to smoke” item. We did not include sleep problems in our analysis since this item was not expected to change during the smoking sessions. Exhaled breath carbon monoxide level (eCO), a biomarker of smoke exposure, was assessed using a handheld monitor (Smokerlyzer Micro, Bedfont Scientific) at time 0 and 5 min to determine eCO boost (eCO at time 5 min minus eCO at time 0).

After each smoking session, measures of subjective effects were completed. The 11-item modified Cigarette Evaluation Questionnaire (mCEQ)^33 34^ includes five subscales: Smoking Satisfaction, Psychological Reward, Aversion, Enjoyment of Respiratory Tract Sensations, and Craving Reduction, with items rated from 1 (not at all) to 7 (extremely likely). Five visual analog scale items ranging from 0 (‘not at all’) to 100 (‘extremely’) assessed wanting to smoke the product again, liking the product, enjoying the product, finding the product pleasurable and satisfying.^35 36^ Behavioral intentions to use the menthol cigarette alternatives were also collected. Participants reported how likely they were to: ‘try this product again,’ ‘purchase this product for personal use,’ and ‘use this product regularly’ if menthol cigarettes were no longer available to be purchased. Participants also completed the Cigarette Purchase Task,^37 38^ a behavioral economic task that assesses hypothetical tobacco product consumption across varying prices. Demand indices include demand intensity (*Q*_0_; the number of products consumed per day when free), essential value (EV; a measure of reinforcing efficacy that measures the rate of change in demand elasticity across the range of prices), P_*max*_ (the price associated with the maximal expenditure, i.e., the highest price before the curve changes from inelastic to elastic), and breakpoint (the last price in which consumption is greater than 0), with higher scores indicating greater abuse liability. Finally, following the completion all of smoking sessions, participants selected their most preferred menthol alternative product to completely substitute for their UBMC for one week.

### Data Analytic Plan

The study was powered based on a laboratory study examining the abuse liability of cigarettes containing different doses of nicotine,^39^ such that with 80 participants there was over 80% power to detect decreases in product satisfaction of up to 50% as compared to UBMC and decreases of 68%-72% in the cigarette purchase task indices of maximum expenditure, maximum price and price sensitivity.

Topography measures were winsorized at the 1^st^ and 99^th^ percentiles and analyzed using repeated measures analysis of variance (ANOVA) with Tukey’s adjustment for all pairwise comparisons between products. For QSU, linear mixed effects models with Holms procedure to adjust for post-hoc comparisons were used to assess differences between products. Subjective smoking experiences were analyzed using repeated measures ANOVA models with Tukey’s adjustment while behavioral intentions were modeled with mixed effects multinomial logistic regression. For all models, log transformations were employed as necessary to satisfy assumptions. All analyses were conducted in SAS 9.4 (SAS Institute, Cary NC).

Demand data were fit to the normalized Zero-Bound Model of Demand (ZBEn) using the freely available GraphPad Prism® template provided by the Institute for Behavior Resources (https://ibrinc.org/behavioral-economics-tools/). To assess normality for all demand indices, we conducted the D’Agostino-Pearson omnibus normality test using GraphPad Prism® version 9. Results indicated the distributions for all demand indices deviated from a Gaussian distribution; therefore, we used the non-parametric Friedman test with Dunn’s correction for multiple comparisons for all analyses (see Supplemental Appendix for detailed methods).

## RESULTS

### Participant Demographics and Smoking History

98 participants enrolled in the study, with 80 completing all phase 1 visits and included in the analyzed sample. Participants had a mean age of 37.1 years (SD=7.5), were predominantly female (75.0%), white (72.5%) and non-Hispanic or Latino (93.8%; Table 1). Participants reported smoking an average of 11.3 cigarettes per day (standard deviation [SD]=5.1) and smoking at this frequency for the last 15.4 years (SD=10.2), with a median FTND score of 3 (interquartile range [IQR] 2-5), indicating a moderate level of dependence. Participants reported minimal past 30-day use of other tobacco products (Table 1).

**Table 1.** Demographics and Tobacco Use History (n=80)

|  | Analyzed Sample<br>(n=80) |  |
| --- | --- | --- |
| Demographics |  |  |
| Age (*mean, SD*) | 37.13 | 7.54 |
| What term below best describes your ethnicity? (*n, %*) |  |  |
| Hispanic or Latino | 5 | 6.3 |
| Not Hispanic or Latino | 75 | 93.8 |
| What term(s) below best describe your race? (*n, %*) |  |  |
| Black or African American | 16 | 20.0 |
| White or Caucasian | 58 | 72.5 |
| Bi or Multi-racial | 6 | 7.5 |
| Below is a list of terms that people often use to describe their sexuality or sexual orientation. Please check the term that best applies to you. (*n, %*) |  |  |
| Gay | 3 | 3.8 |
| Bisexual | 14 | 17.5 |
| Straight/Heterosexual | 62 | 77.5 |
| Queer | 1 | 1.3 |

|  | **Analyzed Sample**<br>**(n=80)** |  |
| --- | --- | --- |
| What sex were you assigned at birth (what the doctor put on your birth certificate)? (*n, %*) |  |  |
| Male | 20 | 25.0 |
| Female | 60 | 75.0 |
| What is the highest level of school you have completed? (*n, %*) |  |  |
| 12th grade, no diploma | 1 | 1.3 |
| High school graduate / GED | 13 | 16.3 |
| Some college, no degree / Associates degree | 45 | 56.3 |
| Bachelor's degree / Master's degree | 21 | 26.3 |
| Which of the following categories best describes your total household income in the past 12 months? (*n, %*) |  |  |
| Less than \$35k | 36 | 45.0 |
| \$35k to \$149,999 | 44 | 55.0 |
| On average, about how many cigarettes do you currently smoke EACH DAY? (One pack usually equals 20 cigarettes). (*mean, SD*) | 11.28 | 5.05 |
| Years smoked at this frequency (*mean, SD*) | 15.39 | 10.15 |
| Usual brand of store-bought cigarettes (*n, %*) |  |  |
| Marlboro | 21 | 26.3 |
| Newport | 21 | 26.3 |
| Camel | 20 | 25.0 |
| Maverick | 6 | 7.5 |
| American Spirit | 3 | 3.8 |
| Other | 9 | 11.3 |
| Years smoked this brand of cigarettes (*mean, SD*) | 10.16 | 8.23 |
| Fagerstrom Test for Nicotine Dependence Score (*median, IQR*) | 3 | 2 – 5 |
| **Other Tobacco Product Use in Past 30 Days (*n, %*)** |  |  |
| Any | 14 | 17.5 |
| Pipe (with tobacco, not including hookah) | 1 | 1.3 |
| Cigars (like Cohiba or Romeo y Julieta) | 1 | 1.3 |
| Cigarillos | 1 | 1.3 |
| Little cigars or filtered cigars | 0 | 0.0 |
| E-cigarette or vaping device (like JUUL, blu, Vuse, MarkTen, Suorin) | 13 | 16.3 |
| Smokeless tobacco (like chewing tobacco, snuff, or dip) | 1 | 1.3 |
| Snus (like Camel Snus) | 0 | 0.0 |
| Hookah/shisha/waterpipe/hookah tobacco | 1 | 1.3 |

### Smoking Topography and eCO Boost

Table 2 compares all four products on topography and exposure measures. Compared to smoking UBMC, participants demonstrated a greater flow rate when smoking mRYO; and greater puff duration, total puffing time, and eCO boost when smoking mFLC, as well as a lower flow rate and max puff volume. When smoking NMC, participants had shorter puffing times and smaller average and total puff volumes than any of three mentholated products.

**Table 2.** Smoking Topography for Usual Brand Menthol Cigarettes and Menthol Cigarette Alternatives (n = 80)

|  | Usual brand<br>menthol<br>cigarette<br>(UBMC) | Menthol roll-your-own<br>cigarette<br>(mRYO) | Menthol filtered<br>little cigar<br>(mFLC) | Non-menthol<br>cigarette<br>(NMC) |
| --- | --- | --- | --- | --- |
|  | M (SD) | M (SD) | M (SD) | M (SD) |
| Total Smoking Time (min) | 5.11 (0.35) | 5.06 (0.19) | 5.12 (0.51) | 5.09 (0.47) |
| Total Puffing Time (min) | 0.36 (0.10)<sup>cd</sup> | 0.36 (0.11)<sup>cd</sup> | 0.47 (0.16)<sup>abd</sup> | 0.33 (0.10)<sup>abc</sup> |
| Average Puff Duration (s) | 2.04 (0.57)<sup>c</sup> | 2.07 (0.65)<sup>cd</sup> | 2.65 (0.91)<sup>abd</sup> | 1.88 (0.61)<sup>bc</sup> |
| Average Flow Rate (ml/s) | 23.10 (6.78)<sup>bc</sup> | 24.70 (7.56)<sup>acd</sup> | 16.96 (5.48)<sup>abd</sup> | 22.16 (7.27)<sup>bc</sup> |
| Average Inter-puff Interval (s) | 27.43 (1.83)<sup>c</sup> | 27.51 (1.24)<sup>c</sup> | 26.40 (2.61)<sup>abd</sup> | 27.76 (1.55)<sup>c</sup> |
| Inhaled Volume (mL) | 461.68 (140.54)<sup>d</sup> | 484.54 (127.13)<sup>cd</sup> | 433.07 (128.73)<sup>bd</sup> | 389.42 (120.21)<sup>abc</sup> |
| Average Puff Volume (mL) | 45.58 (13.61)<sup>d</sup> | 48.15 (12.68)<sup>cd</sup> | 42.46 (12.91)<sup>bd</sup> | 39.03 (11.93)<sup>abc</sup> |
| Max Puff Volume (mL) | 63.14 (19.35)<sup>cd</sup> | 64.18 (15.53)<sup>cd</sup> | 56.57 (18.92)<sup>ab</sup> | 56.34 (16.04)<sup>ab</sup> |
| CO Boost (ppm) | 8.15 (4.09)<sup>c</sup> | 7.40 (3.03)<sup>c</sup> | 9.31 (4.40)<sup>abd</sup> | 7.75 (2.91)<sup>c</sup> |

Mean (M) and standard deviations (SD) for topography measures. p-values estimated from repeated measures ANOVA with Tukey adjustment for all pairwise comparisons. All measures were winsorized at the 1<sup>st</sup> and 99<sup>th</sup> percentiles. Superscripts denote differences in pairwise comparisons between study products at the p<0.05 level. <sup>a</sup> Differs from UBMC; <sup>b</sup> Differs from mRYO; <sup>c</sup> Differs from mFLC; <sup>d</sup> Differs from NMC

### Cigarette Craving and Withdrawal

Mean values for QSU-brief desire and relief factors for all four products over time are depicted in Figure 1A and 1B, respectively. Significant within-participant reduction was observed in both subscales for all products following the initial directed puffing segment, supporting the ability of each of the four products to reduce craving. Significant between-group differences comparing UBMC to the study products were not observed for desire or anticipated relief. Similarly, significant within-participant reduction was observed in withdrawal symptoms for all products following the initial directed puffing segment, but no between-group differences were observed (Figure 1C and 1D).

**Figure 1.**
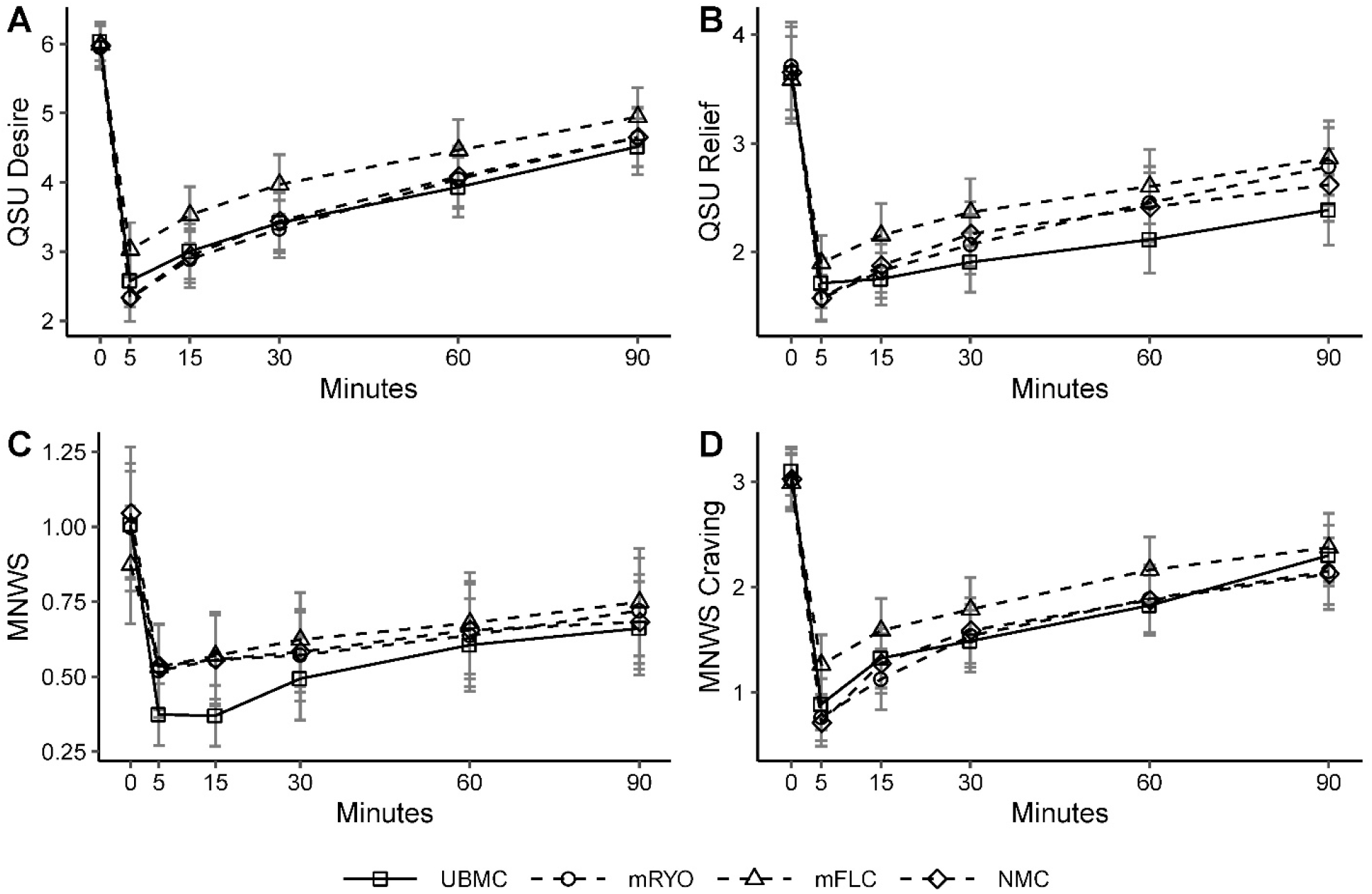
Measures of cigarette craving (Tiffany-Drobes Questionnaire of Smoking Urges; QSU) and withdrawal (Minnesota Nicotine Withdrawal Scale; MNWS) for each product (n = 80) (A) QSU – Desire; (B) QSU – Relief; (C) MNWS withdrawal symptoms; (D) MNWS Craving. Mean and 95% CI estimated immediately before (0) and after the smoking session (5 minutes), and at 15, 30, 60, and 90 minutes.

### Product Demand Indices

Demand curves for four participants (5%) were determined to be non-systematic and removed from analyses. Post-hoc analyses for multiple comparisons indicated significantly greater addiction potential of UBMC when compared to all three alternative products for measures of demand intensity, EV, and breakpoint (p<0.05; Figure 2). Among alternative products, mRYO indicated the greatest addiction potential, with significantly higher intensity and EV than both mFLC and NMC, significantly higher P_*max*_ than NMC, and significantly higher breakpoint than mFLC (p<0.05). There were no significant differences between NMC and mFLC.

**Figure 2.**
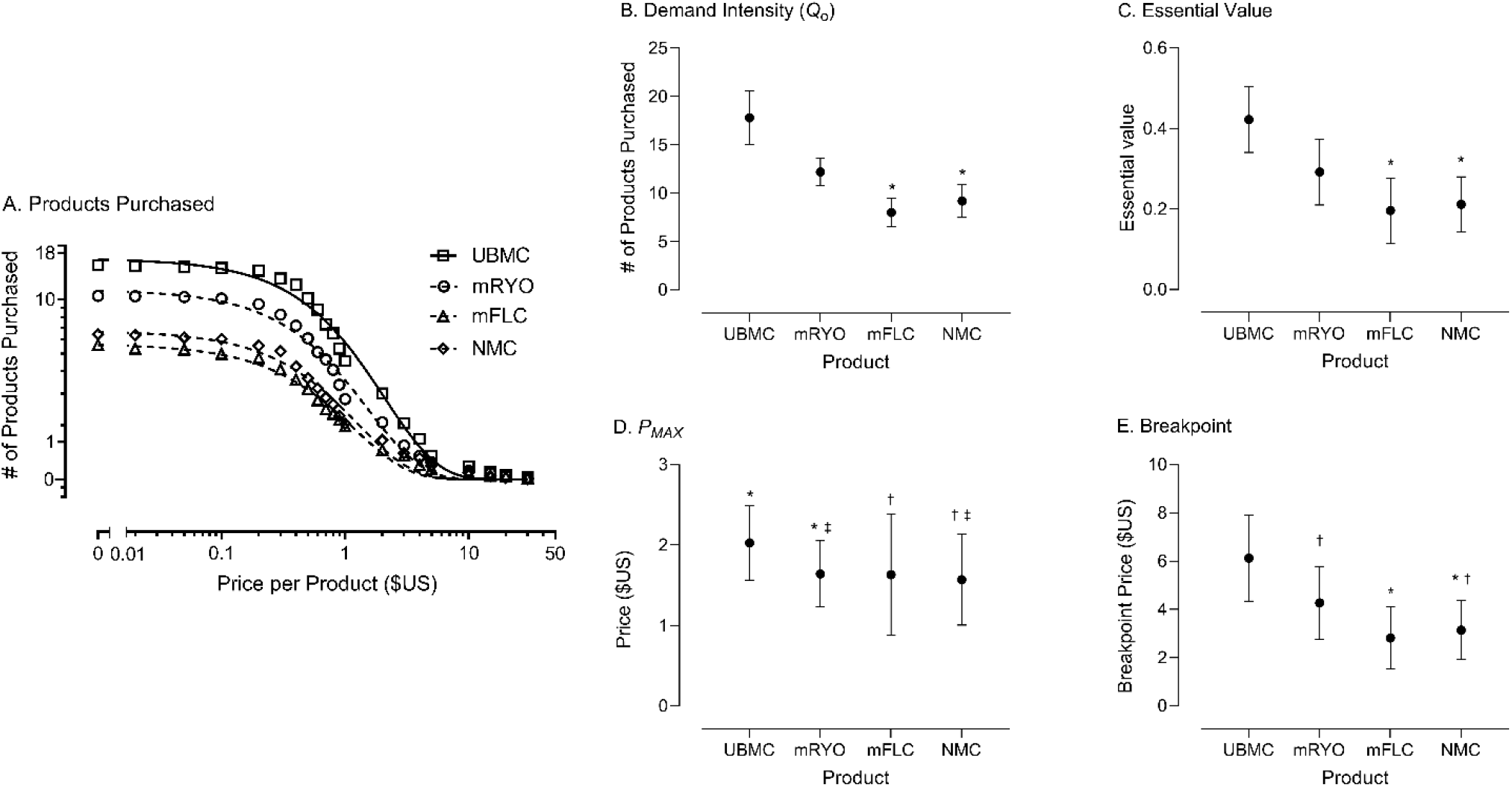
Behavioral economic measures of addiction potential by study product (n=76). (A) Overall demand across all four products. Data points indicate estimated daily consumption (y-axis) across varying price points per cigarette ranging from $0 (free) to $30 (x-axis) for all participants. (B-E) Demand indices across all four products. Data points represent mean scores across participants with 95% CIs. Along the x-axis is product type and along the y-axis are the respective scores or price in $US. For all indices, a higher score or price indicates greater abuse liability. Data points that do not share a symbol differ significantly (p < .05). Note: four participants were removed from analyses due to non-systematic data.

### Subjective Smoking Experience, Behavioral Intentions and Product Selection

Figure 3 depicts mean ratings for all subjective smoking experience items. Compared to UBMC, participants reported significantly lower levels of wanting to smoke the product again, liking, enjoyment, pleasure, and satisfaction for each of the alternative products (p<0.001; Figure 3A). Among the alternative products, participants reported the most favorable subjective experience when smoking mRYO (p<0.001) compared with mFLC and NMC, with no significant differences between mFLC and NMC (Figure 3A). Similarly, on the mCEQ, UBMCs were rated as more satisfying, rewarding, had more enjoyable sensations in the throat and chest, and better craving reduction than the alternative products (p<0.05; Figure 3B). However, UBMC had similar levels of aversion to NMC and higher levels compared to mRYO and mFLC (p<0.05; Figure 3B). Participants were also significantly more likely to want to try again (p<0.001), purchase (p<0.001), and use the mRYO product regularly (p<0.001) compared with mFLC and NMC, with no significant differences between mFLC and NMC (Figure 3C). Consistent with these findings, 65.0% (n = 52) of participants chose mRYO as their preferred menthol alternative to use during phase 2, 22.5% (n = 18) chose NMC, and 12.5% chose mFLC (n = 10).

**Figure 3.**
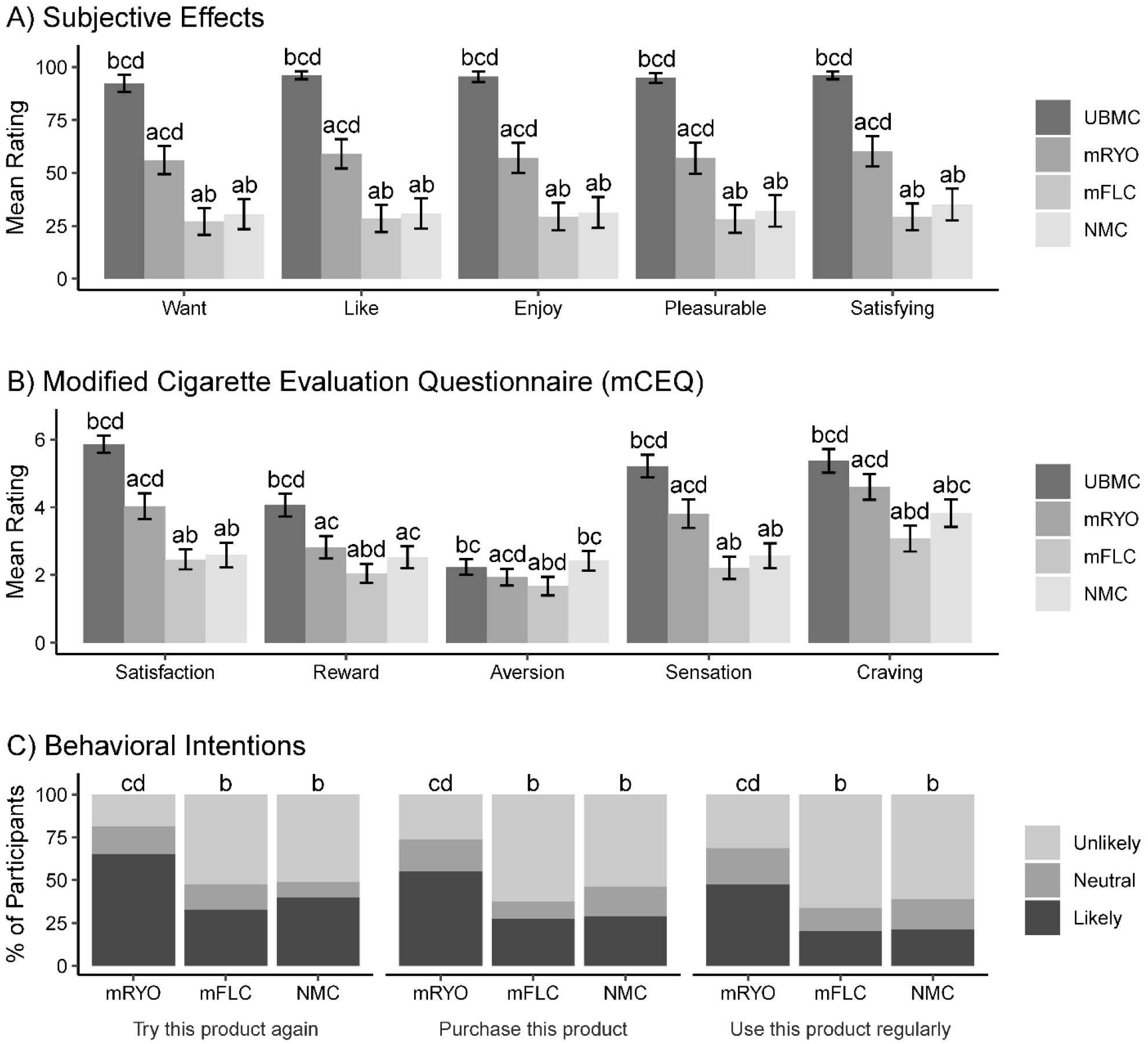
Subjective effects, modified Cigarette Evaluation Questionnaire, and behavioral intentions by study product (n = 80) P-values estimated from mixed effects models accounting for repeated measures; Superscripts denote differences in pairwise comparisons between study products at the p<0.05. ^a^ Differs from UBMC; ^b^ Differs from mRYO; ^c^ Differs from mFLC; ^d^ Differs from NMC.

## DISCUSSION

Using a large within-subjects study of adults who smoke menthol cigarettes and multiple methods of assessing addiction potential, our study expands prior cross-over studies of menthol and non-menthol cigarettes^40-42^ to include other potential menthol cigarette substitutes, specifically menthol-flavored filtered little cigars and the combination of mentholated pipe tobacco and tubes in a menthol roll-your-own cigarette. Study findings were generally consistent with hypotheses that menthol cigarette alternatives would result in similar smoking topography and carbon monoxide (CO) exposure, but fewer positive subjective effects and lower demand. All products suppressed craving and withdrawal, with few differences over time across the four study products. Findings on subjective effects of each product were also similar to other studies,^41 42^ with participants reporting the most favorable subjective effects for their UBMC, with mRYO cigarettes rated next highest and outperforming the other two menthol cigarette alternatives. In line with behavioral intention data on likelihood of trying, purchasing, and using the product regularly, pre-filled mRYO cigarettes were the products chosen by most participants to be used in a subsequent one-week trial at home and the most highly rated cigarette alternative, suggesting their potential appeal as a menthol cigarette substitute. These findings are of particular importance, given the components of this product: pipe tobacco, which now comprises most of the loose tobacco market,^27^ and cigarette tubes, which have been authorized by the FDA in prior substantial equivalence (SE) applications. While the FDA used its enforcement authority in 2013 to call out misbranding of roll-your-own cigarette tobacco as pipe tobacco,^43^ retailer education in 2021 embraced slippage between the product categories and encouraged convenience store owners and operators to promote both to their clients.^44^ Given the 2020 court order vacating the FDA’s health warning requirement for pipe tobacco,^45^ continued ambiguity in differentiating pipe from roll-your-own tobacco, and anticipated FDA action on menthol cigarettes and cigars, our findings suggest that components of menthol roll-your-own products, including menthol rolling papers, cigarette tubes, and pipe tobacco, be considered for inclusion under a menthol cigarette ban.

Beyond measures of the subjective experiences of smoking, topography data can estimate both addiction potential and toxicity of menthol cigarette alternatives. Findings showed that UBMC and mRYO cigarettes were used similarly, with flow rate being the only measure that differed between the two products; this may be related to a lower density of tobacco in the roll-your-own product than the commercial menthol cigarette. There were several topography measures that differed, however, between UBMCs and the other two study products, which may be related to the more negative subjective ratings of these products. Consistent with other studies,^40 41^ participants had shorter puff duration, lower average puff volume, and lower total inhaled smoke volume when using NMC compared with any of the mentholated products. The higher smoke volume seen for our three menthol products may result in higher exposure to nicotine, tobacco-specific nitrosamines, and ultrafine particulates.^40^ Novel findings supported that compared with all other products, mFLC had a higher puffing time, higher puff duration, lower average flow rate and higher CO boost. While speculative, this finding may reflect both the nature and greater density of tobacco in the filtered cigar product.^46^ These data suggest that menthol filtered cigars, used as cigarettes,^47^ may induce greater smoke exposure in their users.

Strengths of our study include use of a within-subjects design, multiple methods of estimating the addiction potential of menthol cigarette alternatives, and a large clinical laboratory sample of adults who currently smoke menthol cigarettes. The fact that our sample had a high proportion of people who identified as white, female, and of lower socioeconomic status is both a strength and a limitation of our study. Even though the prevalence of menthol cigarette use is highest among Black adults who smoke,^3 11^ there remains a larger absolute number of White adults who smoke menthol cigarettes in the U.S. Our sample reflects the midwestern city in which it was recruited, but is likely generalizable to a broader population of menthol cigarette smokers in the U.S., including women and people of lower socioeconomic status who have a higher prevalence of menthol cigarette use.^3 11^ Our study’s use of a limited number of products to evaluate menthol cigarette alternatives does not reflect the range of alternative products that could be substituted for menthol cigarettes under a potential ban, but recent research using an online experimental tobacco marketplace to simulate product choice following a menthol cigarette ban supports menthol little cigars, non-menthol cigarettes, menthol cigarillos, and menthol vapes as potential substitutes.^48^ Expanding our multi-method design to a broader range of products may identify the most likely menthol cigarette alternative; our current findings suggest that menthol pipe tobacco and tubes should be a target for research and regulation.

## Data Availability

The corresponding author will make deidentified participant data and the data dictionary available following publication. Institutions and individuals wishing to access any resources or data must contact the corresponding author. Data will only be made available to those whose proposed use of the data has been approved by the corresponding author. Data will be made available for the sole purpose of replicating the analyses reported in the manuscript. The recipient must agree to not transfer the data to other users and that the data are only to be used for research purposes. The PIs will require requestors of data to sign a data sharing agreement that will ensure (1) Use of the data are only for research purposes (2) Data security using appropriate technology/firewalls, (3) Destruction of data after data analysis and (4) Proper citation in publications or other written materials. A record of transfer of data and a copy of the dataset that was distributed will be kept by The Ohio State University.

## Contributors

TW and AV conceived of and designed the study. TM managed the study and was overseen by TW and AV. AH, JS, and TE conducted and are responsible for the data analysis. TW and AV wrote the manuscript and TM, AH, JS, TE, and JT reviewed, edited and approved the final version. TW, AV, and AH had full access to all the data in the study and take responsibility for the integrity of the data and the accuracy of the data analysis.

## Ethics Approval

Ethics approval was obtained from the Ohio State University Institutional Review Board (IRB Protocol Number: 2019C0107). The trial was registered in clinicaltrials.gov (NCT04844762).

