## Supplemental Appendix for "Addiction Potential of Combustible Menthol Cigarette Alternatives: A Randomized Crossover Trial"

#### SUPPLEMENTARY APPENDIX

Addiction and Behavior Related to Menthol Cigarette Substitutes

Principal Investigator: Theodore L. Wagener, Ph.D., Ohio State Comprehensive Cancer Center

Principal Investigator: Andrea Villanti, Ph.D., University of Vermont Dept. of Psychiatry

### Abstract

Recent changes in population patterns of tobacco use in youth and adults underscore the potential substitution of other menthol tobacco products for cigarettes in the face of a ban on menthol in cigarettes. The increased use of cigars and pipe tobacco in youth following the 2009 ban on flavored cigarettes is particularly important given that flavored filtered little cigars are often indistinguishable from cigarettes and flavored pipe tobacco can be used to make roll-your-own cigarettes. Mentholated pipe tobacco (for roll-your-own cigarettes, mRYO), menthol filtered little cigars (mFLC), and non-menthol cigarettes (nmC) appear to be plausible substitutes for menthol cigarettes. The goal of the current study is to examine the abuse liability and substitutability of these potential menthol cigarette substitutes using an in-laboratory and ad libitum outpatient mixed design. One hundred current menthol cigarette smokers (n=50 aged 18-24 years, n=50 aged 25+ years) will complete a three-phase study: in Phase 1, utilizing a randomized crossover design, participants' will complete 4 smoking sessions, smoking a different product each session to examine each product's abuse liability, demand, and topography. Products will include participants' usual brand menthol cigarette (UBMC) as well as 3 commercially-available alternatives, including mFLC, an mRYO product, and non-mentholated cigarette (nmC). Participants will complete one-week of daily diaries after smoking session 1 in phase 1 to assess their usage of usual brand menthol cigarette (UBMC). In Phase 2, to assess uptake, changes in subjective effects, and use over time, participants will select their preferred study product from Phase 1 and instructed to completely substitute the product for their UBMC for one week. Participants will complete daily diaries during this period to more accurately assess substitution and perceived effects in real-time. In Phase 3, participants will complete a final in-lab visit to assess the substitutability of their preferred product from Phases 1 and 2, under simulated ban conditions using a progressive ratio task. In all phases, multiple domains of abuse liability will be assessed, including product administration (in-lab topography and daily self-report measures), product liking/craving and withdrawal suppression (in-lab and daily diary self-report). This study will be the first to estimate the substitutability of potential menthol cigarettes substitutes in adult smokers, which could impact the health benefit of a public health standard banning menthol in cigarettes. It will evaluate characteristics and perceptions (e.g., satisfaction, taste) of these products that may increase the likelihood of substitutability. Findings from this study will provide key information on the potential unintended consequences of a ban on menthol in cigarettes (i.e., the extent to which these substitutes would appeal to and be used by existing menthol cigarette smokers). They may also inform how FDA treats other non-cigarette tobacco products used as menthol cigarette substitutes in future proposed rulemaking, for example, extending the ban to menthol filtered little cigars or menthol pipe tobacco.

<u>Project Narrative/relevance to public health:</u>
FDA has repeatedly indicated its intent to pursue a ban on menthol in cigarettes. However, estimating the impact of a potential ban on menthol in cigarettes requires accounting for likely substitutes in the marketplace that may also need to be restricted to effectively protect public health. The proposed study uses an in-laboratory and *ad libitum* outpatient mixed design to examine the abuse liability and substitutability of plausible menthol cigarette alternatives, including menthol filtered cigars, menthol roll-your-own cigarettes, and non-menthol cigarettes, in a sample of current adult menthol cigarette smokers.

Addiction and Behavior Related to Menthol Cigarette Substitutes

### A. Specific Aims

While the prevalence of cigarette smoking in the U.S. has continued to decrease,[7] the proportion of menthol cigarette users increased significantly from 35% in 2008-2010 to 39% in 2012-2014.[5] Menthol cigarettes are associated with increased youth smoking initiation, increased nicotine dependence, and decreased adult cessation.[6] Menthol smokers are also more likely to be of low socioeconomic status, female, black or Hispanic, and identify as LGBT compared to non-menthol smokers.[8] As these studies highlight, menthol is strongly associated with facilitating the initiation and maintenance of cigarette smoking, particularly among vulnerable populations.

The 2009 Family Smoking Prevention and Tobacco Control Act banned characterizing flavors in cigarettes and their components. Tobacco companies, however, exploited loopholes in this regulation to maintain sales of their products. For example, Djarum, previously a clove flavored *cigarette*, launched clove filtered *cigars* in anticipation of the ban, and sales of their clove filtered cigars increased by more than 1400% between 2009 and 2012.[9] Moreover, menthol was not included in the ban, and the use of menthol cigarettes as well as flavored cigars and pipe tobacco increased in youth following the flavored cigarette ban, suggesting substitution of other available flavored tobacco products.[10] Thus, while manufactured flavored cigarettes are no longer available, the public health benefit of this product standard was attenuated by the presence of accessible commercially-available substitutes in the marketplace.

For any proposed regulatory action, the FDA must estimate the range of potential impacts on behavior and health. Estimating the impact of a potential ban on menthol in cigarettes, therefore, requires accounting for likely substitutes in the marketplace that may also need to be restricted to effectively protect public health. Current tobacco products, including menthol filtered little cigars (mFLC), menthol roll-your-own (mRYO) tobacco and cigarette tubes, and non-menthol cigarettes (nmC), are relevant targets.[11]

The goal of the proposed study is to examine the abuse liability and substitutability of plausible menthol cigarette alternatives. Using an in-laboratory and *ad libitum* outpatient mixed design, current menthol cigarette smokers (N=80) will complete a three phase, 3-week study: in *Phase 1,* utilizing a randomized crossover design, participants will complete [*4 smoking sessions*], smoking a different product each session to examine each product's abuse liability, demand, and topography. Products will include participants' usual brand menthol cigarette (UBMC) as well as 3 commercially-available alternatives, including mFLC, mRYO product, [*and non-mentholated cigarette (nmC)*.] Participants will complete one-week of daily diaries after smoking session 1 in phase 1 to assess their usage of usual brand menthol cigarette (UBMC). In *Phase 2*, to assess uptake, changes in subjective effects, and use over time, participants will [*select their preferred study product from Phase 1*] and be instructed to completely substitute the product for their UBMC for one week. Participants will complete daily diaries during this period to more accurately assess substitution and perceived effects in real-time. In *Phase 3,* participants will complete a final in-lab visit to assess the substitutability of [*their preferred product from Phases 1 and 2*,] under simulated ban conditions using a progressive ratio task. In all phases, multiple domains of abuse liability will be assessed, including product administration (in-lab topography and daily diary self-report measures), product liking/craving and withdrawal suppression (in-lab and daily diary self-report).

Aim 1: To assess the abuse liability of menthol cigarette alternatives. H1a: Alternatives will [have similar use topography and significantly reduce nicotine craving/withdrawal similar to UBMC, but (H1b) UBMC will show significantly greater demand indices and liking/satisfaction compared to alternatives. H1c: Among alternatives, nmC will show the greatest demand and liking/satisfaction, followed by mFLC, and lastly mRYO.

Aim 2: To assess the substitutability of menthol cigarette alternatives. H2a: A significantly higher portion of product preference selections for Phase 2 will favor nmC than all other alternatives. H2b: Participants' use of study products will significantly increase over the one-week substitution period. H2c: Under simulated UBMC ban conditions, >80% of participants will substitute at least 50% of their UBMC use with study product.

Aim 3: To evaluate which product characteristics and perceived effects influence greater substitution. H3: Participants reporting higher product satisfaction, including "throat hit", menthol-specific sensory effects, craving reduction, improved mood with use, and ease of use [on daily diaries, will report the greatest substitution of study product for UBMC.

### B. Significance

**Menthol cigarette prevalence is increasing:** In the face of historic declines in the prevalence of cigarette smoking in the U.S.,[7] the proportion of menthol cigarette users increased significantly from 35% in 2008-2010 to 39% in 2012-2014.[5] Significant increases in menthol cigarette prevalence occurred in all age groups, with youth (12-17 years old) and young adult (18-24 years old) smokers reporting the highest prevalence of menthol use among smokers (53.9% and 50.0%, respectively).[5] These findings were echoed by data from the 2013-2014 Population Assessment of Tobacco and Health (PATH) Study, with 60% of youth and 47% of young adult smokers using mentholated cigarettes.[4] These changes are consistent with growth in menthol cigarette market share[12-14] and menthol cigarette prevalence can only be expected to increase in youth and young adults given recent expansions in the distribution of menthol cigarettes by the largest U.S. cigarette companies.[15-18]

**Role of menthol in smoking initiation and maintenance:** Reviews of tobacco industry documents underscore the relationship between menthol cigarette use, youth smoking initiation and tobacco dependence, as understood and manipulated by the tobacco industry.[19-21] The appeal of menthol flavoring has been demonstrated to influence intention to smoke and initial smoking,[22,23] with youth more likely to experiment with menthol cigarettes than older age groups.[3,5,24,25] Additionally, young smokers who start with menthol cigarettes are more likely to increase or maintain their smoking behavior over time.[26,27] There are several mechanisms by which menthol in cigarettes has demonstrated to influence the initiation and maintenance of smoking, as indicated by tobacco industry documents as well as independent research: 1) menthol's cooling and analgesic properties mask the harshness and taste of cigarette smoke, making it more appealing; 2) menthol's refreshing sensory qualities increase the positive, or rewarding, properties associated with smoking[28,29]; 3) menthol inhibits nicotine metabolism, causing the smoker greater systemic exposure to nicotine[30]; and 4) menthol may change smokers' puff topography causing them to take more puffs.[31] These mechanisms, long known by the tobacco industry, allowed them to engineer a nicotine delivery device that would not only attract new smokers but also make it more difficult for established smokers to quit.

**Menthol cigarettes, public health, and threats to the efficacy of a ban on menthol cigarettes:** The 2009 Family Smoking Prevention and Tobacco Control Act banned certain characterizing flavors in cigarettes and their components (i.e., tobacco, filter, and paper). The law did not include menthol, nor did it address flavors in non-cigarette tobacco products.[32] However, the Act makes clear that the Food and Drug Administration (FDA) has the authority to issue a product standard to ban menthol in cigarettes, or any other tobacco product, to protect public health. Reviews of the scientific evidence by the FDA and its Tobacco Product Scientific Advisory Committee led to a report concluding that it is "likely that menthol cigarettes pose a public health risk above that seen with non-menthol cigarettes" [33] and "removal of menthol cigarettes from the marketplace would benefit public health in the United States."[34] FDA has continued to request information on the potential effects of a ban on menthol in cigarettes, including in the July 2017 announcement of its comprehensive approach to tobacco and nicotine regulation.[35] The 2009 ban on flavored cigarettes provides a key example of the potential intended and unintended consequences of such a ban. A recent study using data from the 1999-2013 National Youth Tobacco Surveys showed that the flavored cigarette ban was associated with reductions in the prevalence of past 30-day cigarette smoking and number of cigarettes smoked in youth, as intended.[10] However, youth prevalence of past 30-day flavored cigar, pipe, and menthol cigarette use increased following the 2009 ban, suggesting substitution of other flavored tobacco products for flavored cigarettes.[10] This is consistent with evidence of tobacco companies exploiting loopholes in tobacco regulation to maintain sales of their flavored products.[36,37] Djarum, for example, launched clove filtered cigars in the U.S. in anticipation of the 2009 ban on flavored cigarettes and sales of their clove filtered cigars increased by more than 1400% between 2009 and 2012.[9] Thus, while manufactured flavored cigarettes are no longer available, the public health benefit of this product standard was attenuated by the presence of accessible commercially-available substitutes in the marketplace.

Recent trend data highlight growth in U.S. sales of mentholated products, including filtered cigars from 2011 to 2015.[14] Population data show significant correlation between cigar use and menthol cigarette use.[5] These patterns of co-use may be related to the pervasiveness of characterizing flavors, including menthol, in these products.[38,39] Studies on effects of a hypothetical ban on menthol in cigarettes among menthol smokers support behavioral intentions to switch to another tobacco product[40,41] or to non-menthol cigarettes.[41,42] Importantly, these data suggest that poly-use of menthol tobacco products is already occurring, possibly enabling future substitution of other tobacco products for menthol cigarettes in response to an FDA ban. They also highlight non-menthol cigarettes as a possible substitute under such a ban.

**Implications:** Data on recent changes in the tobacco marketplace and population patterns of tobacco use in youth and adults underscore the potential substitution of other tobacco products for menthol cigarettes in the context of a ban. The increased use of cigars and pipe tobacco in youth following the 2009 ban on flavored cigarettes is particularly important given that flavored filtered cigars are often indistinguishable from cigarettes and flavored pipe tobacco is used to make roll-your-own cigarettes. Mentholated pipe tobacco (for RYO cigarettes), menthol filtered cigars, and non-menthol cigarettes appear to be plausible substitutes for menthol cigarettes. The goal of the current study is to examine the abuse liability and substitutability of these potential menthol cigarette substitutes in the laboratory and in an extended observation period. Findings from this study will provide key information on the potential unintended consequences of a ban on menthol in cigarettes (i.e., the extent to which these substitutes would appeal to and be used by existing menthol cigarette smokers). They may also inform how FDA treats other non-cigarette tobacco products used as menthol cigarette substitutes in future proposed rulemaking, [for example, extending the ban to menthol filtered little cigars or menthol pipe tobacco. The scientific premise of this study is that examining abuse liability and substitutability of menthol cigarette substitutes will inform estimates of the public health impact of a ban on menthol in cigarettes. This information is required for FDA's regulatory impact analysis to pursue such a ban. This study will also provide a model for future research on the potential public health impact of flavor bans in non-cigarette products, noted as another priority by FDA in their plan for comprehensive nicotine regulation.[35]

### C. Approach and Preliminary Studies

Our team brings combined expertise in all areas necessary for the successful implementation of a clinical/clinical laboratory study examining the substitutability of other combusted mentholated non-cigarette tobacco products for menthol cigarettes. Specifically, our team has strong expertise conducting survey and longitudinal cohort research examining the 1) the role of menthol in smoking initiation, dependence, and cessation,[3,5,6,8,12,27,51,52,55-57,59,97] 2) the impact of a menthol ban on population health including the substitutability of non-menthol and menthol cigarettes,[6,51,52,57 and 3]) the use of flavored and non-cigarette tobacco products among youth, young adults, adults.[3,4,60,65] We have also conducted five clinical/human laboratory studies examining the pharmacological, toxicological, and physiological effects of non-cigarette tobacco products including two ongoing studies examining the impact of flavors and sweeteners on waterpipe tobacco smoking (R03DA041928; R03DA041928-02S1). In each of these studies, we assessed the abuse liability, topography, nicotine delivery, toxicant exposure, and use behaviors of the non-cigarette tobacco product under investigation. All of these methods are very familiar to our team and we have experienced great success utilizing them. Our team has also conducted several short-term as well as long-term randomized trials assessing tobacco use behavior.[72-83] Lastly, our team has significant experience to collect daily diary data to examine tobacco use and behavioral correlates among smokers.[98-105] These studies have resulted in excellent retention (>80%) with high-rates of daily diary completion (80-88%).

***Theoretical Framework:*** The proposed study is guided generally by behavioral economic theory[1] as well as an established framework for abuse liability assessment used by the FDA to assess drug products, including tobacco.[2] Specifically, abuse liability assessment involves determining the likelihood that use of a product will lead to persistent and problematic use through a series of specific tests across multiple domains (**Figure 1**) including: 1) self-administration tasks, to determine a product's rate of self-administration over time; 2) forced choice tasks, to determine if one product is preferred over another; 3) hypothetical purchase tasks, to determine how much of a product is consumed at different prices or changes in consumption relative to changes in the price of a different product; and 4) positive and negative subjective effects, to determine the psychoactive effects of a drug; and 5) suppression of withdrawal/craving with product use, to determine whether a product can prevent the effects of abstinence in nicotine dependent individuals. Products with a higher abuse liability will be self-administered at a relatively higher rate, preferred over the other product when forced to choose, less sensitive to higher costs to obtain it, greater positive effects/fewer side effects, and more effectively suppress craving and withdrawal symptoms. The proposed study will examine each of these domains.

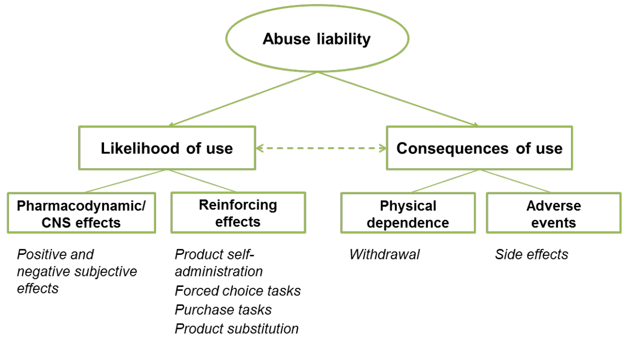

**Figure 1.** Abuse liability model and constructs (adapted from Carter et. al, 2009)

### D. Research Design and Methods

#### 1. Design Overview

**Figure 2.** Study Overview

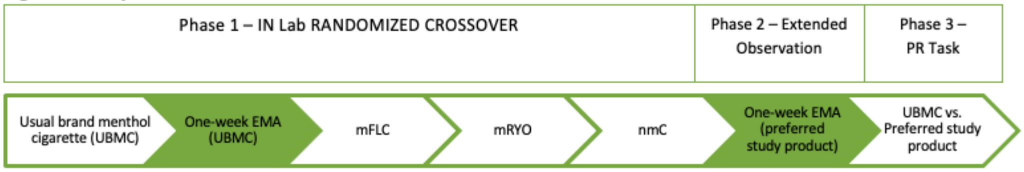

**Figure 2** depicts the sequence of the proposed study. Using an in-laboratory and *ad libitum* outpatient mixed design, 80 current menthol cigarette smokers will complete a three phase, 3-week study. Over the 3-week study time period, participants will attend 5 in-person lab visits. At the beginning of each visit, pregnancy tests will be completed to ensure that the participant is not pregnant. During visit 1, participants will complete a PROP taste test to measure a participants' perceived intensity to taste. In *Phase 1*, utilizing a randomized crossover design, participants will complete 4 smoking sessions, each session smoking a different product examining each participants' puff topography while sampling the product, the product's ability to suppress nicotine craving and withdrawal, and the product's demand indices. Products will include participants' usual brand menthol cigarette (UBMC) as well as 3 commercially-available alternatives, including an mFLC, a pre-assembled mRYO product (menthol tobacco + menthol tube), and an nmC. All sessions will occur following 12-hrs of nicotine abstinence and be separated by 48-hrs. Participants will complete daily diaries for one-week during this period to more accurately assess current use of their UBMC. In *Phase 2*, participants will *select their preferred study product from Phase 1* and be instructed to completely substitute the product for their UBMC for one week. Participants will complete daily diaries during this

period to more accurately assess degree of substitution and perceived effects in real-time. In *Phase 3,* participants will complete a final in-lab visit to assess the substitutability of their preferred product, under simulated ban conditions using a progressive ratio task. In all phases, multiple domains of abuse liability will be assessed, including product administration (in-lab puff topography and daily diary self-report measures), product liking and craving and withdrawal suppression (in-lab and daily diary self-report), and hypothetical purchase tasks to simulate demand.

#### 2. Study Procedures

Enrolled menthol smokers will complete four, 2-hr long counterbalanced smoking sessions that are preceded by 12 hours of overnight tobacco abstinence, biochemically verified by exhaled carbon monoxide (eCO$\leq$10ppm). Sessions will be separated by a standard 48-hour washout period. Participants will smoke one cigarette ad libitum and a puff topography device will discretely record smoking behavior throughout the session, including puff duration, number of puffs, puff volume, and time between puffs. eCO, and self-report measures of craving and withdrawal will be recorded immediately before and at several times after the smoking session. Self-report measures of abuse liability will also be completed during the sessions. During the preferred study product observational use period, participants will be provided a weeks' worth of a potential menthol substitute at no cost. Consistent with our previous switching studies, study products will be provided in a 1.2 to 1 ratio based on self-reported use at the time of screening. This slight over appropriation helps to ensure that participants have enough study product available so to not artificially limit use, while also not oversupplying and potentially artificially increasing use behind what would be usual. Participants will be instructed to completely switch and exclusively use the study product during the 1-week time period. During this period, participants will complete daily diary once a day. Following the one-week use period, participants will come to the lab following 12-hr nicotine abstinence confirmed by eCO. To simulate the effect that restricting menthol in cigarettes would have on increasing (or not) preference for other alternative menthol substitutes, participants will complete a 90-minute concurrent choice task with differential cost (effort) required to earn 2 puffs from their UBMC (10 clicks increasing to 7200) versus their preferred study product (always 10 clicks) from Phase 2. This session will last approximately 3 hours.

<u>Recruitment Feasibility and Retention</u>

<u>Recruitment:</u> Based on our team's previous studies we conservatively assume a 20% attrition rate; thus, we will need to recruit 100 participants to have 80 complete the study. We are confident that our recruitment approaches will yield sufficient numbers given our successful history of recruitment for other tobacco-related research of similar design.[75,76,82] Menthol cigarette smokers will be recruited from advertisements through a variety of media outlets and the internet, including Study Search, as well as community events. Participants from other studies who have agreed to be contacted regarding other study opportunities will also be contacted. Staff from those studies will prepare contact letters/emails and call participants on behalf of this study. Participants interested will be referred to this study for screening. Participants will access the screening questionnaire using a public survey link generated by REDCap. Participants who meet the following eligibility criteria will be asked to take part in the study.

<u>Inclusion Criteria:</u> 1) a current menthol cigarette smoker (>90% menthol cigarette use; ≥5 cigarettes per day) for at least the past 6 months, 2) between 21-24 (young adult or 25-50 years old (aged 25+), 3) willing to provide informed consent and abstain from all tobacco and nicotine use for at least 12 hours prior to the five lab sessions, 4) willing to complete two weeks of daily dairies, 5) read and speak English, and 6) have access to a smart phone or email

Exclusion Criteria: 1) self-reported diagnosis of lung disease including asthma, cystic fibrosis, or chronic obstructive pulmonary disease, 2) history of cardiac event or distress within the past 3 months, 3) currently pregnant, planning to become pregnant, or breastfeeding (will be verified with urine pregnancy test), 4) use of other tobacco products (e.g., e-cig, cigar, etc.) >5 days in the past month, 5) current marijuana use >5 days per month, 6) any use of other illicit drugs during the last 30 days, 7) currently engaging in a smoking cessation attempt, and 8) currently using one of the alternative menthol study products, and 9) reside in the same household as a participant currently active (have not completed all study visits) in the study. If a participant is ineligible for the study at the time of screening, the participant can be reassessed at a later time to determine if they are now eligible i.e. A participant meets all other eligibility criteria, but is not eligible because they are currently pregnant or breastfeeding. The participant can submit a new screener after they are no longer pregnant or breastfeeding and can be reassessed for eligibility. Reassessment for eligibility will vary based on previous ineligibility criteria and will be determined on a participant by participant basis.

Participants' eligibility will be determined over the phone or via REDCap's online screener. Those who are eligible and willing to participate will be invited to sign an informed consent and complete their baseline visit in a private participant room at the Ohio State University. All participants will be given adequate time to review the informed consent with a trained research staff to help answer any questions that may arise during the consent process. Additionally, a copy of the informed consent will be given to all participants. A pregnancy test will be completed at the initial visit as well as before starting all the in-lab visits to ensure that the participant is not pregnant.

Retention: All participants will receive $50 per completed in-laboratory session, $7 for parking (when applicable), $50 bonus for completing all sessions (up to $335), and up to $30 for completing daily diaries over the two week ad-lib and observational use periods ($30 for 12 more diaries completed, $25 for 11 diaries completed, $20 for 9-10 diaries completed, $15 for 7-8 diaries completed and $0 for 1-6 diaries completed) for a total up to $365. Consistent with our previous studies, payments will be made using the Greenphire ClinCard to increase accountability and facilitate ease of payment. We will also facilitate study calls/visits by offering evening and weekend appointments as well as additional retention strategies (e.g., multiple sources of contact, reminder calls/texts/emails). Participants will receive reminder calls in addition to email or text reminders. Reminders will be sent by text or email based on a participant's preferred method of contact.

**Study Products:** Mentholated pipe tobacco in a roll-your-own cigarette tube (mRYO), menthol filtered little cigars (mFLC), and non-menthol cigarettes (nmC) were chosen as plausible menthol cigarette substitutes that are currently available on the commercial market. To produce the mRYO product, study staff will inject 1 gram of menthol OHM pipe tobacco (www.smokersoutletonline.com/ohm-pipe-tobacco-1-lb.html) into manufactured Hot Rod King (84 mm; www.smokersoutletonline.com/hot-rod-tubes.html) tubes using an electric rolling machine, as described in online user forums. The menthol filtered cigar will be Cheyenne (Cheyennecigars.com) Seneca (senecacigars.com). The nmC will be Newport Non-Menthol Red. All products (mFLC, mRYO, nmC) will be provided to participants in plain boxes without brand or identifying information for the in-lab and *ad libitum* sessions; the box will include a study product ID number sticker for tracking purposes. Brand or identifying information may be present on the actual product.

Detailed Study Procedures

UBMC and Study Product Lab Sessions (Phase 1): Upon arrival at the lab, 12-hr tobacco abstinence will be assessed via self-report and confirmed with exhaled carbon monoxide testing (eCO≤10ppm). Pregnancy exclusion will also be confirmed with a urine test; and breath alcohol concentration (BrAC) tests will ensure BrAC <.01. Participants will not be allowed to eat or drink (other than water) during the session. Participants will be instructed to smoke one UBMC or study product *ad libitum* to completion through a puff topography device. eCO will be collected immediately pre- and post-smoking. Measures of subjective effects (see Table 1) will be collected immediately before the onset of puffing and at 1-min, 15-min, 30-min, 60-min, and 90-min after completion of the cigarette. The Cigarette Purchase Task will also be completed during the lab sessions. Sessions will last approximately 2 hours each. During this period, participants will complete one week of daily diaries to assess their usage of UBMC.

Preferred Study Product *Ad libitum*-Observational Use Period (Phase 2): During the preferred study product observational use period, participants will be provided products at no cost. Study products will be provided in a 1.2 to 1 ratio based on self-reported use at the time of screening. This slight over appropriation helps to ensure that participants have enough study product available so to not artificially limit use, while also not oversupplying and potentially artificially increasing use beyond what would be usual. Participants will be instructed to completely switch and exclusively use the study product during the 1-week time period. During this period, participants will complete one week of daily diaries. If the participant has not responded after 3 prompts, the assessment will be recorded as missed.

Final Lab Session – Simulated Menthol Cigarette Ban Using a Progressive Ratio Task (Phase 3): Following the one-week use period, participants will come to the lab following 12-hr nicotine abstinence confirmed by eCO. To simulate the effect that restricting menthol in cigarettes would have on increasing (or not) preference for other alternative menthol substitutes, participants will complete a 90-minute concurrent choice task with differential cost (effort) required to earn 2 puffs from their UBMC (10 clicks increasing to 7200 ) versus their preferred study product (always 10 clicks) from Phase 2. This session will last approximately 3 hours.

**Data Management**

All data collection will follow HIPAA guidelines. Data will be collected directly from the participant by a research assistant. Data will include participant responses to computer-based and phone-based survey Questionnaires, as well as exhaled carbon monoxide samples and progressive ratio task and puff topography readouts.

Access to Identifiable Information and Data Storage: Only research assistants who have completed training in the ethical conduct of research and the study MPIs (Drs. Wagener and Villanti) will have access to individually identifiable private information about human subjects. All data will be treated as confidential and will never be stored or reported in association with identifying information. Hard copies of signed informed consent and the patients cover sheet which includes contact information will be stored in locked filing cabinets separate from participants' study-related data. A common identification number will link identifiable forms (consent forms and contact information) and study-related data. Computer entered data will be de-identified and password-protected.

**Quality Assurance**

All research staff will have completed Human Subjects and HIPAA training. Standard operating procedures (SOP) have been developed for similar studies run by our lab; we will spend the first month developing the SOP for this protocol. All staff will be trained to ensure adherence to the SOP. As is standard practice for our team's current studies, each visit will have its own checklist of specific measures to be completed and the order in which they are to be administered. On-site personnel will meet face-to-face weekly throughout the study, with Dr. Villanti joining all weekly meetings via Skype.

#### 3. Measures

Questionnaire data will be collected over the phone or in-person by a trained research assistant and data will be entered into a secured and encrypted database using REDCap. See Table 1 for timing of measures. Sociodemographic measures will assess participant age, sex, marital status, ethnicity, employment status, occupation, years of education, and socioeconomic status. Tobacco use history will assess years of smoking, age of smoking onset, average number of cigarettes per day, number and recency of previous 24-hour quit attempts, number of smokers in the household, prior use of nicotine replacement therapy and other stop smoking medications, and history of receiving smoking cessation counseling. It will also assess tobacco type, brand, frequency, quantity, and duration of use all of nicotine/tobacco products including cigars, cigarillos, little cigars, pipe tobacco, chewing tobacco, snuff, snus, EC/vape/mod/APV/e-hookah, combusted tobacco hookah, and dissolvable tobacco. Cigarette Dependence will be measured with the 12-item Cigarette Dependence Scale.[118] Exhaled carbon monoxide will be assessed at the start of each study visit. Abuse liability of products will be measured across several domains 1) smoking puff topography, 2) subjective effects, 3) behavioral economic choice tasks, and 4) craving for and suppression of craving and withdrawal. Smoking puff topography will be measured using the eTOP which uses a pressure transducer integrated into a plastic cigarette holder to produce measures of puff count, puff duration, inter-puff-interval, puff flow rate, average puff volume, and total puff volume. Puff topography is a validated and sensitive behavioral measure of abuse liability, is highly stable and associated with level of dependence and predicts level of exposure to harmful tobacco-related toxicants.[119-121] An adapted version of the Drug Effects/Liking Questionnaire[122] will assess the desire and liking of UBMC and all three study products, positive and negative effects (i.e., side effects), and perceived strength and effectiveness. The modified Cigarette Evaluation Questionnaire (mCEQ) will also assess subjective responses to cigarettes (e.g., reward, satisfaction).[123,124] The Cigarette Purchase Task[125,126] will ask participants how much they would be willing to pay (ranging from 0¢ to $1,120) to smoke each product. Given that the study products will look similar to cigarettes, we will retain the original language (e.g., "1 cigarette") in the purchase task. Willingness to spend more will indicate greater abuse liability. Smoking urges/craving will be measured using the Tiffany-Drobes Questionnaire of Smoking Urges: Brief Form.[127] This is a 10-item measure where participants rate smoking-related items ("All I want right now is a cigarette.") on a 7-point Likert scale ranging from 'strongly agree' to 'strongly disagree'. Similar to previous studies, we will collapse the items into two previously identified factors (Factor 1: strong desire and intention to smoke; Factor 2: anticipation of relief from withdrawal symptoms). Nicotine withdrawal will be assessed using the empirically validated 15-item version of the Minnesota Nicotine Withdrawal Scale.[128] This measure assesses smoking craving, anger/irritability, anxiety, depressed mood, restlessness/difficulty concentrating, increased appetite, sleep problems, and somatic symptoms (nausea, constipation, sore throat, dizziness, coughing). Subjective effects (daily diary) of the Phase 2 substitute product will be derived from daily diaries assessing product satisfaction and pleasure. Substitutability of products will be assessed using a Cross-Price Task, a Progressive Ratio (PR) Task and daily diaries. A Cross-Price Task in Phases 1 and 3 will estimate substitutability of the study product for the UBMC.[129,130] Participants will be asked how many study products and UBMCs they would consume when the price of the study product is fixed at $1 and the UBMC prices escalate. The data are then fit to an exponential equation that indicates whether the fixed-price product substitutes for the primary product, and the degree of substitution. Cross price elasticity (CPE) for each study product compared to UBMC > 0.2 indicates substitution, CPE < -0.2 indicates complementarity, and CPE between -0.2 and 0.2 indicate independence of the two products.[131] Consistent with previous studies conducted by Dr. Tidey,[89,93,94,132,133] the PR task will simulate the effect that restricting menthol in cigarettes would have on increasing (or not) preference for other alternative menthol substitutes. Participants will complete a 90-minute concurrent choice task with differential cost (effort) required to earn the reinforcement (2 puffs) from their UBMC and the study product (mFLC, mRYO or nmC). Puffs from the study product can be earned by clicking a computer mouse 10 times on a picture of the study product, but to earn two puffs of the UBMC, they will be required to make escalating response requirements (computer mouse clicks) according to the following schedule: 10, 160, 320,

640, 1280, 2400, 3600, 4800, 6000, 7200. A maximum of 10 reinforcers (20 puffs) per session will be allowed. The proportion of reinforcers earned is considered to provide an index of the strength of the reinforcing effects of the product. Participants will be informed of the differential sequence between products and instructed that the sessions are 3 hours in length no matter how much or how little they respond. Daily diaries will assess UBMC/study product smoked per day, product satisfaction and pleasure (see Table 1 for specific daily diary measures). Substitution assessed via use behavior during Phase 2 will be operationalized as the ratio of study product to UBMC used, with a ratio > 0 indicating any substitution and a ratio > 1 indicating substitution of study product for the UBMC at least 50% of the time.

Table 1. Measures

| **Measures** | **Phase 1:** | | | **Phase 2:** | **Phase 3:** |
|---|---|---|---|---|---|
| | **3 in-lab visits** | | | **1-week** | ***Final Lab*** |
| | Visit 1 | Visit 2 | Visit 3 | Visit 4 | Visit 5 |
| ***Background Measures*** | | | | | |
| Exhaled CO Abstinence verification | X | X | X | X | X |
| Pregnancy Test | X | X | X | X | X |
| Exhaled carbon monoxide | X | X | X | X | X |
| Breath Alcohol Concentration (BrAC) | X | X | X | X | X |
| Taste testing strips (PROP) | X | | | | |
| Sociodemographic measures | X | | | | |
| Cigarettes Use/Tobacco Use Hx (EDSHC) | X | | | | |
| Menthol Subscales | X | | | | |
| Readiness Rulers | X | | | | |
| Product Use Status | X | | | | |
| Product Use Detailed Assessment | X | | | | |
| Fagerstrom | X | | | | |
| ***Abuse liability*** | | | | | |
| Smoking topography | X | X | X | X | |
| Drug Effects/Liking Questionnaire | X | X | X | X | X |
| Modified Cigarette Evaluation Questionnaire (mCEQ) | X | X | X | X | X |
| Tiffany-Drobes Questionnaire of Smoking Urges: Brief Form (modified) | X | X | X | X | X |
| MNWS | X | X | X | X | X |
| Cigarette Purchase Task | X | X | X | X | X |
| Subjective effects | X | X | X | X | X |
| ***Substitutability*** | | | | | |
| Cross Price Elasticity Task | | X | X | X | X |
| Progressive ratio task (UBMC vs. study product - computer task) | | | | | X |
| Record final product selected | | | | X | |

Addiction and Behavior Related to Menthol Cigarette Substitutes

| | | | | |
|---|---|---|---|---|
| Use behavior (Daily Diary) | X | | | X |
| **Daily diary** | | | | |
| · # Study products smoked | | | | X |
| · # Non-study products smoked | X | | | X |
| · Other tobacco use | X | | | X |
| Modified Cigarette Evaluation Questionnaire (mCEQ) | X | | | X |
| Drug Effects/Liking Questionnaire | X | | | X |
| Behavioral Intentions | | | | X |
| Tiffany-Drobes Questionnaire of Smoking Urges: Brief Form (modified) | X | | | X |
| MNWS | X | | | X |

### E. Statistical Methods

#### 1. Power Analysis

Since our main goal is to evaluate the substitutability of menthol alternatives in the context of a menthol cigarette ban, statistical power is based on differences in abuse liability measures (Hypothesis 1a) and product substitution (Hypothesis 2c). Sample size estimates relied on means and standard deviations of data collected as part of a University of Vermont laboratory study examining the abuse liability of cigarettes containing different doses of nicotine.[89] A sample size of 80 subjects has 81% power to detect decreases in product satisfaction of up to 50% compared to their usual brand (mean for UBMC 5.5, SD 1.3). In addition, a sample size of 80 participants has greater than 80% power to detect decreases of 68%-72% in the cigarette purchase task indices of maximum expenditure, maximum price and price sensitivity. This sample size also provides 81% power to estimate 80% use of the preferred alternate product more than half the time during Phase 2 with a 95% confidence interval of 66%-94%.

#### 2. Data Analytic Plan

Statistical analyses will be performed using SAS 9.4 and an alpha of .05. Background measures will be summarized by product (UBMC, mFLC, mRYO, nmC), as appropriate. Continuous variables will be presented as mean ± SD; categorical variables will be presented as counts and proportions. We will apply a transformation to normalize the distribution and stabilize the variance of the residuals where appropriate.

Hypotheses 1a and 1c: We will use Repeated Measures Analysis of Variance with Tukey-adjustment to examine differences between the alternatives and the UBMC in abuse liability assessed at each product lab session. Assuming statistically significant product differences, this post-hoc test will allow us to not only compare each product to the UBMC, but will also allow comparison of the alternative products with each other. Because product introduction in Phase 1 will be randomized using a Latin Square, a fixed effect for session and a random effect for sequence will be included in all analyses.

Addiction and Behavior Related to Menthol Cigarette Substitutes

Hypothesis 2a: Product preference will be examined using a Chi-Square Test, comparing the proportions of participants choosing each alternate product for further use.

Hypothesis 2b: We will examine the trajectory of use behavior outcomes using Linear Mixed Model (LMM) regression analysis. We will employ a random intercept or slope parameter, as appropriate, and model the covariance structure for the repeated outcome measures, while accounting for potential confounders, including gender and baseline cigarettes/day.

Hypothesis 2c: This aim will be analyzed as a Chi-Square Test of Goodness of Fit to test the proportion of use compared to 80%.

Hypothesis 3: We will examine whether differences in product characteristics and changes in perceived effects collected via daily diary are associated with increased substitution of the alternate product over the seven-day period using generalized LMM regression analysis, similar to that outlined in H2b.

Exploratory analyses regarding moderation: For both lab session measures in phase 1 and product substitution in phase 2, we will examine whether age moderates the differences in abuse liability measures and perceived effects of the abuse liability on use behavior outcomes by including an age-by-predictor effect in all analyses. Age will be dichotomized at less than 25 years of age compared to age 25 or older to conform to the recruitment strategy.

##### 3. Missing Data

In the event of missing data, we will contact participants immediately or censor at the point of loss if they cannot be contacted. If the combined missing rate is very small (<5%) and the data are confirmed to be missing at random, then we may safely perform the data analysis on the available data using maximum likelihood procedures. If the missing rate is high, then we will explore sequential multiple imputation (SMI).

#### F. Gender/Minority/Pediatric Inclusion for Research

##### 1. Inclusion of Women and Minorities

According to 2018 US Census estimates, 51.3% of Columbus residents are female. We expect that the proportion of female participants will likely be somewhat larger given our previous studies with smokers (55-62% female). According to 2018 US Census estimates, the racial composition of individuals living in Columbus is 60.5% White, 28.3% Black or African American, 5.2% Asian, 0.2% American Indian/Alaska Native, 0.0% Native Hawaiian/Other Pacific Islander, and 4.1% two or more races. The ethnic composition of individuals living in Columbus is 6.0% Hispanic/Latino and 94.0% Non-Hispanic/Latino. We expect that our distribution will be similar to these but may potentially have a larger distribution of ethnic and racial minorities, given our previous studies and that menthol smokers tend to more often be black or Hispanic. However, we will continuously monitor enrollment in order to ensure that we are meeting recruitment goals to avoid under-recruiting minorities. If the targeted enrollment for minorities is not met because they do not respond to the advertisements, we will make special efforts to solicit their participation by advertising in community newspapers, local church organizations, and community centers.

##### 2. Inclusion of Children

Participation in the proposed study will be restricted to individuals 21 to 50 years of age. This exclusion is for two primary reasons: 1) the use of tobacco products by minors is illegal, and 2) the concern of introducing and potentially addicting children and adolescents to another tobacco product.

### G. Human Participants

#### 1. Recruitment and Informed Consent

At first contact, all participants will be screened according to the study's inclusion/exclusion criteria. Those who are eligible will be given a brief verbal overview of the study and invited to participate. Informed consent (including a description of the nature, purpose, risks, and benefits of the study) will take place through both oral and written explanation of the study. The voluntary nature of the study and the participant's right to withdraw at any time will be stressed during the consent process; a copy of the informed consent will be provided to the participant in written form at the time of consent for them to keep. Informed consent will be collected by IRB approved study personnel. Recruitment script and materials, consent forms and all study procedures will be approved by the OSU Institutional Review Board. All participants will provide written consent before any study data is collected.

#### 1. Potential Risks and Protections Against Risk

There are minimal risks associated with this protocol. The protocol requires menthol smokers 21 to 50 years of age and older to undergo 12 hours of tobacco/nicotine abstinence on six occasions and to attempt to substitute another combusted mentholated nicotine product for their current combusted mentholated nicotine product. Tobacco/nicotine abstinence can lead to withdrawal symptoms that include irritability, anxiety, restlessness, hunger, and difficulty sleeping.[126] The effects can be uncomfortable but are not dangerous. Risks and side effects related to the cigarette products that are commercially available include:

- Nicotine addiction: Nicotine is a highly addictive chemical found in cigarettes, and toxic at certain doses. It negatively affects the brain, nervous system and heart, and excessive exposure can result in poisoning, particularly in young children and pets. It also causes blood vessels to contract, increasing your blood pressure and pulse rates.
- Chronic diseases including COPD, bronchitis, emphysema, coronary heart disease, stroke and cancer.
- Smoking can also cause infertility and peptic ulcer disease, as well as slow the healing of wounds. It's the leading cause of preventable illness and death in the U.S.

The risk associated with substituting one combustible menthol product for another is also low. It is very unlikely that there is any difference in the level of harm between the participants' usual brand and the study products; therefore, substitution is unlikely to increase participants' exposure to harmful constituents over their usual brand. We are also attempting to mitigate the risk of artificially increased use due to receiving product by only giving participants study products in a 1.2 to 1 ratio.

We will withdraw participants who become pregnant, begin to breastfeed or receive diagnosis for a cardiovascular disease during the course of the study.

The risk of undermining smoking cessation is also potential risk; however, we will only recruit smokers not currently engaged in a smoking cessation attempt, and we will provide all participants at the end of the study with a referral to the Ohio Tobacco Quit Line (1-800-QUIT-NOW).

Protection against loss of confidentiality and privacy will be maintained by numerically coding all data, disguising identifying information, and keeping data locked in file drawers or in a secure, password-protected database. Only study research assistants and the PI will have the information that connects participant's name and ID number. All electronic data will be numerically coded and stored in a password protected database, on a password protected computer in a secure research space. Participant information will be accessible only to research staff, who are pledged to confidentiality and complete training in the ethical conduct of research (i.e., both HIPAA and CITI trainings). Identifying information will not be reported in any publication.

##### 2. Potential Benefits of the Proposed Research

Whereas no assurance can be made to an individual participant that s/he will personally benefit from this research, the experience should be beneficial. The immediate benefits of this research are scientific in nature, which in the long-term should benefit society as a whole. The study will also benefit menthol smokers as a group by providing information as to the abuse liability of other mentholated products; and serve as evidence to inform regulatory action that improves public health. Overall, it is expected that the potential benefits to participants in the proposed study outweigh the potential risks.

#### H. Data and Safety Monitoring Plan

Data will be analyzed initially after 20 participants are accrued, to ensure electronic data capture systems employed (i.e., REDCap) are accurately capturing data and to ensure the format and completeness of all data collected.

##### 1. Adverse events

Adverse events will be assessed by study staff at each follow-up visit via participant self-report and managed immediately. All adverse events will be reported to the OSU IRB. We will monitor for risk of smoking by screening participants for general medical precautions (pregnancy, cardiovascular disease). Any adverse events, breaks of confidentiality, or any other data or safety issues that arise will be discussed immediately between study personnel and Dr. Wagener. Dr. Wagener will be responsible for completing an Adverse Events Form should an event occur. Dr. Wagener will report Serious Adverse Events to the OSU IRB within 24 hours of having received notice of the event. Dr. Wagener will gather any information needed to investigate the event and to determine subsequent action. Any subsequent action will be documented and reported to the OSU IRB and the Program Officer at NIH. Adverse event reports will be reviewed annually with the OSU IRB to ensure participant safety.

**Collection of Adverse Events**

The collection of adverse events will be on a self-report basis and logged within an electronic data capture system (REDCap) or collected using standardized paper forms and will only be identified with the study's ID of the participant.

Addiction and Behavior Related to Menthol Cigarette Substitutes

### Addendum

#### COVID-19 Related Procedures

Due to the COVID-19 pandemic, processes and procedures have been implemented to help protect participants and research staff. These processes and procedures are to be followed as long as social distancing requirements are necessary for conducting study visits.

Only one study participant per study coordinator will attend study visits at the CTR at any given time. All study participants will be provided with a face mask upon entry. Only one coordinator will meet the participant at their car for a temperature check, direct the participant into the building, and the two of them will ride the elevator to the 4$^{th}$ floor physically distanced at least 6 ft apart, both wearing masks. No more than 2 persons may ride the elevator at any given time. The participant will be immediately escorted to a private exam/draw room. Therefore, there will be no waiting in open lobby/waiting areas.

When in the exam room, the study coordinator will stand at least 6 feet away from the study participant to give instructions. Afterwards, the study coordinator will leave the exam room to allow the study participant to conduct the instructed procedures. The study coordinator and study participant will be at least 6 feet away from one another and wearing protective masks at all times during each visit.

Each study coordinator will have a designated exam/draw room and smoking room in which to conduct their designated research study. Each smoking room is separated from the staff control station in the hallway by its own door and contains a large window for the study coordinator to be able to see in and monitor study participant activity within the room. There is also a speaker and microphone system within each smoking room along with the Genetech software system on the outside of each room at the smoking room computer stations. Therefore, the study coordinator and study participant can communicate without being in the room together.

For study measures which cannot be physically distanced, appropriate PPE will be worn at all times by research staff during these procedures including goggles, face masks, gloves, and isolation gowns or lab coats.

After each participant visit is complete, there will be at least a 45-minute period for cleaning and air exchanges in the negative pressure rooms and for cleaning exam rooms and equipment before the next participant visit. All smoking rooms are under negative pressure with a ventilation rate of 36.8 – 44.1 air changes per hour (ACH).

Addiction and Behavior Related to Menthol Cigarette Substitutes

15084646.

31. Yerger VB, McCandless PM. Menthol sensory qualities and smoking topography: a review of tobacco industry documents. *Tob Control.* 2011; 20 Suppl 2:ii37-43. PubMed PMID: 21504930. PubMed Central PMCID: 3088462.
32. Fagan P, Pohkrel P, Herzog T, Pagano I, Vallone D, Trinidad DR, Sakuma KL, Sterling K, Fryer CS, Moolchan E. Comparisons of three nicotine dependence scales in a multiethnic sample of young adult menthol and non-menthol smokers. *Drug Alcohol Depend.* 2015; 149:203-211. PubMed PMID: 25744873.
33. U.S. Food and Drug Administration. *Preliminary Scientific Evaluation of the Possible Public Health Effects of Menthol versus Nonmenthol Cigarettes.* Silver Spring, MD: Center for Tobacco Products, Food and Drug Administration; 2013.
34. Tobacco Products Scientific Advisory Committee. *Menthol Cigarettes and Public Health: Review of the Scientific Evidence and Recommendations.* Rockville, MD: Center for Tobacco Products, Food and Drug Administration; 2011.
35. U.S. Food and Drug Administration. FDA's Plan for Tobacco and Nicotine Regulation. 2017; https://www.fda.gov/TobaccoProducts/NewsEvents/ucm568425.htm. Accessed February 7, 2018.
36. Delnevo CD, Giovenco DP, Miller Lo EJ. Changes in the Mass-merchandise Cigar Market since the Tobacco Control Act. *Tob Regul Sci.* 2017; 3(2 Suppl 1):S8-S16. PubMed PMID: 28317004. PubMed Central PMCID: PMC5351883.
37. Delnevo CD, Hrywna M, Giovenco DP, Miller Lo EJ, O'Connor RJ. Close, but no cigar: certain cigars are pseudo-cigarettes designed to evade regulation. *Tob Control.* 2017; 26(3):349-354. PubMed PMID: 27220622. PubMed Central PMCID: PMC5482568.
38. Delnevo CD, Wackowski OA, Giovenco DP, Manderski MT, Hrywna M, Ling PM. Examining market trends in the United States smokeless tobacco use: 2005-2011. *Tob Control.* 2014; 23(2):107-112. PubMed PMID: 23117999. PubMed Central PMCID: PMC3604094.
39. Delnevo CD, Giovenco DP, Ambrose BK, Corey CG, Conway KP. Preference for flavoured cigar brands among youth, young adults and adults in the USA. *Tob Control.* 2015; 24(4):389-394. PubMed PMID: 24721967.
40. Wackowski OA, Delnevo CD, Pearson JL. Switching to E-Cigarettes in the Event of a Menthol Cigarette Ban. *Nicotine Tob Res.* 2015; 17(10):1286-1287. PubMed PMID: 25634935. PubMed Central PMCID: PMC4592340.
41. Wackowski OA, Manderski MT, Delnevo CD. Young adults' behavioral intentions surrounding a potential menthol cigarette ban. *Nicotine Tob Res.* 2014; 16(6):876-880. PubMed PMID: 24514070. PubMed Central PMCID: PMC4015098.
42. Pearson JL, Abrams DB, Niaura RS, Richardson A, Vallone DM. A ban on menthol cigarettes: impact on public opinion and smokers' intention to quit. *Am J Public Health.* 2012; 102(11):e107-114. PubMed PMID: 22994173.
43. Niaura RS, Villanti AC. Tobacco Use. In: Brown BB, Prinstein MJ, eds. *Encyclopedia of Adolescence.* Vol 3. San Diego: Academic Press; 2011:331-337.
44. Villanti A, Boulay M, Juon HS. Peer, parent and media influences on adolescent smoking by developmental stage. *Addict Behav.* 2011; 36(1- 2):133-136. PubMed PMID: 20855170.
45. Rath JM, Villanti AC, Abrams DB, Vallone DM. Patterns of tobacco use and dual use in US young adults: the missing link between youth prevention and adult cessation. *J Environ Public Health.* 2012; 2012:679134. PubMed PMID: 22666279. PubMed Central PMCID: 3361253.

60. Villanti AC, Richardson A, Vallone DM, Rath JM. Flavored tobacco product use among U.S. young adults. *Am J Prev Med.* 2013; 44(4):388-391. PubMed PMID: 23498105.
61. Richardson A, Ganz O, Pearson J, Celcis N, Vallone D, Villanti AC. How the industry is marketing menthol cigarettes: the audience, the message and the medium. *Tob Control.* 2015; 24(6):594-600. PubMed PMID: 25178275.
62. Villanti AC, Abrams DB, Delnevo CD. Informing policy through tobacco regulatory science: An evolving process. *Health Behavior & Policy Review.* 2014; 1(2):97-102.
63. Villanti AC, Cantrell J, Pearson JL, Vallone DM, Rath JM. Perceptions and perceived impact of graphic cigarette health warning labels on smoking behavior among U.S. Young adults. *Nicotine Tob Res.* 2014; 16(4):469-477. PubMed PMID: 24212476. PubMed Central PMCID: 3954425.
64. Villanti AC, Pearson JL, Cantrell J, Vallone DM, Rath JM. Patterns of combustible tobacco use in U.S. young adults and potential response to graphic cigarette health warning labels. *Addictive Behaviors.* 2015; 42:119-125. PubMed PMID: 25437268.
65. Ambrose BK, Day HR, Rostron B, Conway KP, Borek N, Hyland A, Villanti AC. Flavored Tobacco Product Use Among US Youth Aged 12-17 Years, 2013-2014. *JAMA.* 2015; 314(17):1871-1873. PubMed PMID: 26502219.
66. Delnevo CD, Giovenco DP, Steinberg MB, Villanti AC, Pearson JL, Niaura RS, Abrams DB. Patterns of Electronic Cigarette Use Among Adults in the United States. *Nicotine Tob Res.* 2016; 18(5):715-719. PubMed PMID: 26525063.
67. Glasser A, Johnson A, Rath JM, Williams V, Vallone DM, Villanti AC. Tobacco Product Brand Preference among U.S. Young Adults, 2011-2014. *Tob Reg Sci.* 2015; 2(1):44-55.
68. Glasser AM, Cobb CO, Teplitskaya L, Ganz O, Katz L, Rose SW, Feirman S, Villanti AC. Electronic nicotine delivery devices, and their impact on health and patterns of tobacco use: a systematic review protocol. *BMJ Open.* 2015; 5(4):e007688. PubMed PMID: 25926149. PubMed Central PMCID: 4420972.
69. Kirchner TR, Villanti AC, Tacelosky M, Anesetti-Rothermel A, Gao H, Pearson JL, Ganz O, Cantrell J, Vallone DM, Abrams DB. National enforcement of the FSPTCA at Point-of-Sale. *Tob Reg Sci.* 2015; 1(1):24-35.
70. Kirchner TR, Villanti AC, Cantrell J, Anesetti-Rothermel A, Ganz O, Conway KP, Vallone DM, Abrams DB. Tobacco retail outlet advertising practices and proximity to schools, parks and public housing affect Synar underage sales violations in Washington, DC. *Tob Control.* 2015; 24(e1):e52-58. PubMed PMID: 24570101.
71. D'Silva J, Cohn AM, Johnson AL, Villanti AC. Differences in Subjective Experiences to First Use of Menthol and Non-Menthol Cigarettes in a National Sample of Young Adult Cigarette Smokers. *Nicotine Tob Res.* 2017. PubMed PMID: 29059351.
72. Busch AM, Leavens EL, Wagener TL, Buckley ML, Tooley EM. Prevalence, Reasons for Use, and Risk Perception of Electronic Cigarettes Among Post-Acute Coronary Syndrome Smokers. *J Cardiopulm Rehabil Prev.* 2016. PubMed PMID: 27120039.
73. James SA, Meier EM, Wagener TL, Smith KM, Neas BR, Beebe LA. E-Cigarettes for Immediate Smoking Substitution in Women Diagnosed with Cervical Dysplasia and Associated Disorders. *Int J Environ Res Public Health.* 2016; 13(3). PubMed PMID: 26959042. PubMed Central PMCID: PMC4808951.
74. Leavens EL, Brett EI, Frank S, Shaikh RA, Leffingwell TR, Croff JM, Wagener TL. Association between breath alcohol concentration and waterpipe lounge patrons' carbon monoxide exposure: A field investigation. *Drug Alcohol Depend.* 2017; 170:152-155. PubMed PMID: 27918950.
75. Leavens EL, Driskill LM, Molina N, Eissenberg T, Shihadeh A, Brett EI, Floyd E, Wagener TL. Comparison of a preferred versus non-preferred waterpipe tobacco flavour:

subjective experience, smoking behaviour and toxicant exposure. *Tob Control.* 2017. PubMed PMID: 28381414.

76. Lechner WV, Meier E, Wiener JL, Grant DM, Gilmore J, Judah MR, Mills AC, Wagener TL. The comparative efficacy of first- versus second-generation electronic cigarettes in reducing symptoms of nicotine withdrawal. *Addiction.* 2015; 110(5):862-867. PubMed PMID: 25639148.
77. Lechner WV, Tackett AP, Grant DM, Tahirkheli NN, Driskill LM, Wagener TL. Effects of duration of electronic cigarette use. *Nicotine Tob Res.* 2015; 17(2):180-185. PubMed PMID: 24827788. PubMed Central PMCID: PMC4830219.
78. Meier EM, Tackett AP, Miller MB, Grant DM, Wagener TL. Which nicotine products are gateways to regular use? First-tried tobacco and current use in college students. *Am J Prev Med.* 2015; 48(1 Suppl 1):S86-93. PubMed PMID: 25528714.
79. Tackett AP, Lechner WV, Meier E, Grant DM, Driskill LM, Tahirkheli NN, Wagener TL. Biochemically verified smoking cessation and vaping beliefs among vape store customers. *Addiction.* 2015; 110(5):868-874. PubMed PMID: 25675943.
80. Wagener TL, Beebe, L., Gillaspy, S., Vickerman, K., Carpenter, M., Frank, S. Applying the Risk Reduction Continuum to Smoking Substitution: Results of an Adaptive E-cigarette Switching Trial. Annual INBRE Conference; 2016; North Dakota.
81. Wagener TL, Floyd EL, Stepanov I, Driskill LM, Frank SG, Meier E, Leavens EL, Tackett AP, Molina N, Queimado L. Have combustible cigarettes met their match? The nicotine delivery profiles and harmful constituent exposures of second-generation and third-generation electronic cigarette users. *Tob Control.* 2017; 26(e1):e23-e28. PubMed PMID: 27729564.
82. Wagener TL, Meier E, Hale JJ, Oliver ER, Warner ML, Driskill LM, Gillaspy SR, Siegel MB, Foster S. Pilot investigation of changes in readiness and confidence to quit smoking after E-cigarette experimentation and 1 week of use. *Nicotine Tob Res.* 2014; 16(1):108114. PubMed PMID: 24154511.
83. Wagener TL, Tackett AP, Borrelli B. Caregivers' interest in using smokeless tobacco products: Novel methods that may reduce children's exposure to secondhand smoke. *J Health Psychol.* 2015. PubMed PMID: 25845835.
84. Wagener TL, Floyd, E., Stepanov, I., Driskill, L., Meier, E., Leavens, E., Frank, S., Tackett, A., Molina, N., Queimado, L. Evaluation of Nicotine, Carbon Monoxide, and Total NNAL in Cigarette Smokers and Second and Third Generation E-cigarette Users. *Tobacco Control.* Under review.
85. Tidey JW, Rohsenow DJ, Kaplan GB, Swift RM. Cigarette smoking topography in smokers with schizophrenia and matched non-psychiatric controls. *Drug Alcohol Depend.* 2005; 80(2):259-265. PubMed PMID: 15869844.
86. Murphy JG, MacKillop J, Tidey JW, Brazil LA, Colby SM. Validity of a demand curve measure of nicotine reinforcement with adolescent smokers. *Drug Alcohol Depend.* 2011; 113(2-3):207-214. PubMed PMID: 20832200. PubMed Central PMCID: PMC3025087.
87. MacKillop J, Tidey JW. Cigarette demand and delayed reward discounting in nicotine-dependent individuals with schizophrenia and controls: an initial study. *Psychopharmacology (Berl).* 2011; 216(1):91-99. PubMed PMID: 21327760. PubMed Central PMCID: PMC3640631.
88. Higgins ST, Heil SH, Sigmon SC, Tidey JW, Gaalema DE, Stitzer ML, Durand H, Bunn JY, Priest JS, Arger CA, Miller ME, Bergeria CL, Davis DR, Streck JM, Zvorsky I, Redner R, Vandrey R, Pacek LR. Response to varying the nicotine content of cigarettes in vulnerable populations: an initial experimental

examination of acute effects. *Psychopharmacology (Berl).* 2017; 234(1):89-98. PubMed PMID: 27714427. PubMed Central PMCID: PMC5203959.

Addiction and Behavior Related to Menthol Cigarette Substitutes

100. Businelle MS, Ma P, Kendzor DE, Frank SG, Wetter DW, Vidrine DJ. Using Intensive Longitudinal Data Collected via Mobile Phone to Detect Imminent Lapse in Smokers Undergoing a Scheduled Quit Attempt. *J Med Internet Res.* 2016; 18(10):e275. PubMed PMID: 27751985. PubMed Central PMCID: PMC5088341.
101. Kendzor DE, Businelle MS, Mazas CA, Cofta-Woerpel LM, Reitzel LR, Vidrine JI, Li Y, Costello TJ, Cinciripini PM, Ahluwalia JS, Wetter DW. Pathways between socioeconomic status and modifiable risk factors among African American smokers. *J Behav Med.* 2009; 32(6):545-557. PubMed PMID: 19757014. PubMed Central PMCID: PMC2828046.
102. Kendzor DE, Businelle MS, Poonawalla IB, Cuate EL, Kesh A, Rios DM, Ma P, Balis DS. Financial incentives for abstinence among socioeconomically disadvantaged individuals in smoking cessation treatment. *Am J Public Health.* 2015; 105(6):11981205. PubMed PMID: 25393172.
103. Kendzor DE, Shuval K, Gabriel KP, Businelle MS, Ma P, High RR, Cuate EL, Poonawalla IB, Rios DM, Demark-Wahnefried W, Swartz MD, Wetter DW. Impact of a Mobile Phone Intervention to Reduce Sedentary Behavior in a Community Sample of Adults: A Quasi-Experimental Evaluation. *J Med Internet Res.* 2016; 18(1):e19. PubMed PMID: 26810027. PubMed Central PMCID: PMC4746437.
104. Lam CY, Businelle MS, Aigner CJ, McClure JB, Cofta-Woerpel L, Cinciripini PM, Wetter DW. Individual and combined effects of multiple high-risk triggers on postcessation smoking urge and lapse. *Nicotine Tob Res.* 2014; 16(5):569-575. PubMed PMID: 24323569. PubMed Central PMCID: PMC3977487.
105. Watkins KL, Regan SD, Nguyen N, Businelle MS, Kendzor DE, Lam C, Balis D, Cuevas AG, Cao Y, Reitzel LR. Advancing cessation research by integrating EMA and geospatial methodologies: associations between tobacco retail outlets and real-time smoking urges during a quit attempt. *Nicotine Tob Res.* 2014; 16 Suppl 2:S93-101. PubMed PMID: 24057995. PubMed Central PMCID: PMC3977633.
106. Donny EC, Caggiula AR, Mielke MM, Booth S, Gharib MA, Hoffman A, Maldovan V, Shupenko C, McCallum SE. Nicotine self-administration in rats on a progressive ratio schedule of reinforcement. *Psychopharmacology (Berl).* 1999; 147(2):135-142. PubMed PMID: 10591880.
107. Donny EC, Houtsmuller E, Stitzer ML. Smoking in the absence of nicotine: behavioral, subjective and physiological effects over 11 days. *Addiction.* 2007; 102(2):324-334. PubMed PMID: 17222288.
108. Shiffman S, Hickcox M, Paty JA, Gnys M, Richards T, Kassel JD. Individual differences in the context of smoking lapse episodes. *Addict Behav.* 1997; 22(6):797-811.
109. Wetter DW, McClure JB, Cofta-Woerpel L, Costello TJ, Reitzel LR, Businelle MS, Cinciripini P. A randomized clinical trial of a palmtop computer-delivered treatment for smoking relapse prevention among women. *Psychol Addict Behav.* 2011; 25:365-371.
110. Stone AA, Schwartz JE, Neale JM, Shiffman S, Marco CA, Hickcox M, Paty J, Porter LS, Cruise LJ. A comparison of coping assessed by ecological momentary assessment and retrospective recall. *J Pers Soc Psychol.* 1998; 74(6):1670-1680. PubMed PMID: 9654765.
111. Shiffman S, Paty JA, Gnys M, Kassel JA, Hickcox M. First lapses to smoking: Within-subjects analysis of real-time reports. *J Consult Clin Psychol.* 1996; 64(2):366-379. PubMed PMID: 8871421.
112. Piasecki TM, Cooper ML, Wood PK, Sher KJ, Shiffman S, Heath AC. Dispositional drinking motives: Associations with appraised alcohol effects and alcohol consumption in an

ecological momentary assessment investigation. *Psychol Assess.* 2014; 26(2):363369. PubMed PMID: 24274049. PubMed Central PMCID: Pmc4245013.

113. Simons JS, Dvorak RD, Batien BD, Wray TB. Event-level associations between affect, alcohol intoxication, and acute dependence symptoms: Effects of urgency, self-control, and drinking experience. *Addict Behav.* 2010; 35(12):1045-1053. PubMed PMID: PMC3298685.
114. Swendsen JD, Tennen H, Carney MA, Affleck G, Willard A, Hromi A. Mood and alcohol consumption: An experience sampling test of the self-medication hypothesis. *J Abnorm Psychol.* 2000; 109(2):198204.
115. Kendzor DE, Shuval K, Gabriel KP, Businelle MS, Ma P, High RR, Cuate EL, Poonawalla IB, Rios DM, Demark-Wahnefried W. Impact of a Mobile Phone Intervention to Reduce Sedentary Behavior in a Community Sample of Adults: A Quasi-Experimental Evaluation. *J Med Internet Res.* 2016; 18(1).
116. Businelle MS, Ma P, Kendzor DE, Frank SG, Vidrine DJ, Wetter DW. An Ecological Momentary Intervention for Smoking Cessation: A Feasibility Study. under review.
117. Businelle MS, Ma P, Kendzor DE, Reitzel LR, Chen M, Lam CY, Bernstein I, Wetter DW. Predicting quit attempts among homeless smokers seeking cessation treatment: An ecological momentary assessment study. *Nicotine Tob Res.* 2014; 16:1371-1378. PubMed PMID: 24893602.
118. Etter JF, Le Houezec J, Perneger TV. A self-administered questionnaire to measure dependence on cigarettes: the cigarette dependence scale. *Neuropsychopharmacol.* 2003; 28(2):359-370. PubMed PMID: 12589389.
119. Gass JC, Germeroth LJ, Wray JM, Tiffany ST. The Reliability and Stability of Puff Topography Variables in Non-Daily Smokers Assessed in the Laboratory. *Nicotine*
120. Perkins KA, Karelitz JL, Giedgowd GE, Conklin CA. The reliability of puff topography and subjective responses during ad lib smoking of a single cigarette. *Nicotine Tob Res.* 2012; 14(4):490-494. PubMed PMID: 22039077. PubMed Central PMCID: PMC3313780.
121. Lee EM, Malson JL, Waters AJ, Moolchan ET, Pickworth WB. Smoking topography: reliability and validity in dependent smokers. *Nicotine Tob Res.* 2003;
122. 5(5):673-679. PubMed PMID: 14577984.
123. Zacny JP, Conley K, Marks S. Comparing the subjective, psychomotor and physiological effects of intravenous nalbuphine and morphine in healthy volunteers. *J Pharmacol Exp Ther.* 1997; 280(3):11591169. PubMed PMID: 9067299.
124. Cappelleri JC, Bushmakin AG, Baker CL, Merikle E, Olufade AO, Gilbert DG. Confirmatory factor analyses and reliability of the modified cigarette evaluation questionnaire. *Addict Behav.* 2007; 32(5):912-923. PubMed PMID: 16875787.
125. Westman E, Levin E, Rose J. Smoking while wearing the nicotine patch-is smoking satisfying or harmful. *Clin Res.* 1992; 40(4):A871-A871.
126. Jacobs EA, Bickel WK. Modeling drug consumption in the clinic using simulation procedures: demand for heroin and cigarettes in opioid-dependent outpatients. *Exp Clin Psychopharmacol.* 1999; 7(4):412426. PubMed PMID: 10609976.
127. MacKillop J, Murphy JG, Ray LA, Eisenberg DT, Lisman SA, Lum JK, Wilson DS. Further validation of a cigarette purchase task for assessing the relative reinforcing efficacy of nicotine in college smokers. *Exp Clin Psychopharmacol.* 2008; 16(1):57-65. PubMed PMID: 18266552.
128. Cox LS, Tiffany ST, Christen AG. Evaluation of the brief questionnaire of smoking urges (QSU-brief) in laboratory and clinical settings. *Nicotine Tob Res.* 2001; 3(1):7-16. PubMed PMID: 11260806.
